# Intergenerational transmission of complex traits and the offspring methylome

**DOI:** 10.1101/2024.04.15.24305824

**Authors:** Fiona A. Hagenbeek, René Pool, Austin J. Van Asselt, Erik A. Ehli, August B. Smit, Meike Bartels, Jouke Jan Hottenga, Conor V. Dolan, Jenny van Dongen, Dorret I. Boomsma

**Affiliations:** Department of Biological Psychology, Vrije Universiteit Amsterdam, Amsterdam, the Netherlands; Amsterdam Public Health (APH) research institute, Amsterdam, the Netherlands; Institute for Molecular Medicine Finland (FIMM), HiLIFE, University of Helsinki, Helsinki, Finland; Avera McKennan Hospital, University Health Center, Sioux Falls, SD, USA; Department of Molecular and Cellular Neurobiology, Center for Neurogenomics and Cognitive Research, Amsterdam Neuroscience, Vrije Universiteit Amsterdam, Amsterdam, the Netherlands; Amsterdam Reproduction & Development (AR&D) research institute, Amsterdam, the Netherlands; Department of Complex Trait Genetics, Center for Neurogenomics and Cognitive Research, Amsterdam Neuroscience, Vrije Universiteit Amsterdam, Amsterdam, the Netherlands

**Keywords:** Intergenerational transmission, DNA methylation, polygenic scores, genetic nurture, gene-environment correlation, complex human traits

## Abstract

The genetic makeup of parents can directly or indirectly affect their offspring phenome through genetic transmission or via the environment that is influenced by parental heritable traits. Our understanding of the mechanisms by which indirect genetic effects operate is limited. Here, we hypothesize that one mechanism is via the offspring methylome. To test this hypothesis, polygenic scores (PGSs) for schizophrenia, smoking initiation, educational attainment (EA), social deprivation, body mass index (BMI), and height were analyzed in a cohort of 1,528 offspring and their parents (51.5% boys, mean [*SD*] age = 10 [2.8] years). We modelled parent and offspring PGSs on offspring buccal-DNA methylation, accounting for the own PGS of offspring, and found significant associations between parental PGSs for schizophrenia, EA, BMI, and height, and offspring buccal methylation sites, comprising 16, 2, 1, and 6 sites, respectively (alpha = 2.7 × 10^−5^). More DNA methylation sites were associated with maternal than paternal PGSs, possibly reflecting the maternal pre- and periconceptional environment or stronger maternal involvement in shaping the offspring’s environment during early childhood.

## Introduction

Phenotypic resemblance among parents and their biological offspring can be due to both genetic and environmental influences, with the genetic contribution to phenotypic resemblance manifesting through different sources. The first source applies to all heritable phenotypes: the Mendelian transmission of parental alleles to their offspring, resulting in parents and offspring sharing 50% of their autosomal alleles. A second source is heritable parental behaviors, which shape the environment of the offspring. Here, the effect of parental genotype on offspring phenotypes is mediated by parental traits and behavior, contributing to the offspring environment. This source is denoted “genetic nurture”, because it implies that nurture – the totality of external factors after conception influenced or created by parents – may have a genetic component [1, 2]. Hence, genetic nurture arises from the influence of a heritable parental phenotype on the phenotype of their offspring, which is also known as vertical transmission [3]. Additional forms of indirect genetic effects can encompass other forms of cultural transmission, such as, horizontal transmission between individuals of the same generation, such as siblings [4, 5], and ecological inheritance, involving the transmission of inherited resources and conditions to descendants through niche construction [6].

Several designs can resolve direct genetic effects (the influence of genotype of the offspring on own phenotype) and indirect genetic effects (the above-mentioned genetic nurture). These include multi-generation twin-family designs that infer genetic effects from phenotypic resemblance among family members [7, 8], designs based on measured genetic information, e.g., polygenic scores (PGSs) [1, 2, 9, 10], or designs that combine these two [11]. Through PGS designs, robust evidence for genetic nurture effects in offspring educational attainment (EA) has been found [1, 2, 12].

The mechanism through which direct genetic effects (Mendelian inheritance) contribute to variation in human traits is complex, involving a cascade of transcription and translation of genetic information [13]. Here, we propose to examine DNA methylation as one possible mechanism through which both direct and indirect genetic effects may act. Epigenetic mechanisms such as DNA methylation modulate gene expression [14], but are themselves subject to both genetic [15, 16] and environmental influences [17]. Animal studies have shown that early life environmental exposures like diet, trauma, and social deprivation can induce epigenetic reprogramming in cells. For example, a classic study of rats found that the quality of maternal care early in life leads to epigenetic alterations that affect the behavior of offspring into adulthood [18]. Offspring raised by (non-biological) less-nurturing mothers exhibit increased anxiety as adults, which is attributed to enduring changes in DNA methylation in the brain. This study was based on cross-fostering experiments in which pups born to calm biological dams were raised by anxious adoptive dams, and vice versa, to separate the effect of the environment provided by the mother from genetic transmission [18].

In humans, it is more challenging to disentangle the influence of direct genetic transmission and indirect genetic effects that operate via the environment created by parents. Comprehensive reviews of childhood psychological adversity and DNA methylation indicated that childhood maltreatment and other adversities are associated with differential DNA methylation but called for further study of the impacts of childhood experiences and the effects of genetic transmission of parental psychopathology risk [19–21]. In the present study, we propose an approach to disentangle the influence of direct genetic transmission and indirect genetic effects that operate via the environment created on DNA methylation, by modelling the effects of parental PGSs and offspring PGSs on the offspring DNA methylation profile [9, 10]. Finding an effect of parental PGSs, in addition to the effect of the offspring’s own PGS, would support the hypothesis that genetic nurture acts through the offspring methylome. We obtained parental and offspring PGSs for six traits: schizophrenia, smoking initiation, EA, social deprivation, body mass index (BMI), and height in nuclear twin families. We focused on schizophrenia, EA, BMI, and height because they have high powered genome-wide association studies (GWAS). We focused on smoking due to its well-documented impact on DNA methylation as demonstrated by previous epigenome-wide association studies (EWAS) [22, 23]. Similarly, we examined social deprivation because of its strong theoretical basis suggesting mediation through epigenetic mechanisms [24]. EWAS have revealed widespread associations of DNA methylation and schizophrenia [25, 26], smoking [22, 23], and BMI [27, 28]. PGSs for these traits have been associated with DNA methylation [25, 29–31]. However, to date no studies have been undertaken to separate direct from indirect genetic effects on DNA methylation in offspring, which requires the PGSs of the offspring and the offspring’s parents.

In our study, the epigenetic data were available for 1,528 young twins (51.5% boys, mean [*SD*] age = 10 [2.8] years). DNA samples from buccal cells for genotyping and polygenic scoring were available for the offspring and their parents. DNA methylation was measured by the Illumina Infinium EPIC array, which measures DNA methylation at approximately 850K sites. To reduce multiple testing burden, we selected the top 10% most variable probes (72,889). We have shown in an independent sample of adult Dutch twins that the most variable probes are the most reliable across time, tissue, and platform [32]. DNA methylation levels in offspring were simultaneously regressed on offspring, maternal, and paternal PGSs by linear regression, while correcting for familial clustering.

## Materials and methods

### Study population and procedures

The collection of DNA samples in parents and their offspring was done at home from buccal swabs. Families were recruited from the Netherlands Twin Register (NTR) [33] and the majority took part in the ACTION Biomarker Study, which recruited twin pairs based on their (dis)similarity for childhood aggressive behavior [34, 35]. We first collected data in a pilot study to assess the suitability of the Infinium EPIC array for buccal-derived DNA samples [36] in 96 monozygotic (MZ) twins (47 complete pairs, 56.2% boys, mean [*SD*] (range) age at DNA collection 7.4 [2.4] (1-10) years). Next, 1,141 twins (523 complete pairs, 52.6% boys, mean [*SD*] (range) age at DNA collection = 9.6 [1.9] (5.6-12.9) years) and their parents were included in the main ACTION Biomarker Study [37]. A third dataset included additional twins and siblings in the ACTION Biomarker Study, and unselected MZ twins and triplets from the NTR, totaling 291 children of 1 to 18 years of age (45.7% boys, mean [*SD*] age at DNA collection = 12.1 [4.2] years). DNA methylation data and offspring PGSs were available for all offspring, maternal PGSs were available for 92.1% and paternal PGSs for 81.1% families, with parental PGS of both biological parents available for 78.3% families (**Supplementary Table 1**). All families were of European ancestry.

Parents gave written informed consent for their own and their offspring’s participation. The study was conducted according to the guidelines of the Declaration of Helsinki. The ACTION project was assessed by the Central Ethics Committee on Research Involving Human Subjects of the VU University Medical Center, Amsterdam (NTR 03-180, NTR 25th of May 2007, ACTION 2013/41 and 2014.252), an Institutional Review Board certified by the U.S. Office of Human Research Protections (IRB number IRB00002991 under Federal-wide Assurance FWA00017598).

### Infinium MethylationEPIC BeadChip

DNA methylation in offspring was assessed with the Infinium MethylationEPIC BeadChip Kit, following the manufacturer’s specification (Illumina, San Diego, CA, USA) [38]. A total of 500 ng of genomic DNA from buccal swabs were bisulfite-treated using the ZymoResearch EZ DNA Methylation kit (Zymo Research Corp, Irvine, CA, USA). Datasets 1 and 3 were generated at the Avera McKennan Hospital (Sioux Falls, SD, USA), and dataset 2 at the Human Genotyping Facility (HugeF) of ErasmusMC (Rotterdam, the Netherlands; http://www.glimdna.org/).

As previously described [36, 39], quality control (QC) and normalization of the methylation data were performed with the pipelines developed by the Biobank-based Integrative Omics Study (BIOS) consortium [40]. The probe β-values represent the ratio of the methylated signal intensity to total signal intensity (methylated plus unmethylated) at a given CpG site and can range from 0 (unmethylated) to 1 (fully methylated). Only samples that passed all five quality criteria of MethylAid were retained [41]. In addition, these samples were characterized by correct genetic relationships among the participants (omicsPrint) [42] and the absence of sex mismatches (DNAmArray and meffil) [43]. Functional normalization was performed with the dataset-specific optimum number of principal components. Methylation probes were coded as missing, if they had an intensity value of zero, bead count < 3, or detection p-value > 0.01. A probe was excluded if it overlapped with a SNP or Insertion/Deletion (INDEL), mapped to multiple locations in the genome, or had a success rate < 0.95 across all samples. Sample QC, probe filtering, normalization, and imputation of missing values were done within the three datasets separately. Of 865,859 sites on the array, 789,888 sites, 787,711 sites, and 734,807 sites were retained after QC in datasets 1, 2, and 3, respectively, and we retained the 728,899 overlapping sites. Cellular proportions were predicted in epithelial tissues based on the cell-type deconvolution algorithm Hierarchical Epigenetic Dissection of Intra-Sample-Heterogeneity (HepiDISH) with the reduced partial correlation [44]. After QC, we removed all cross-reactive probes, probes in SNPs or on the X or Y chromosome and then imputed missing methylation β-values (up to 5%; probes with higher missingness were excluded) with the imputePCA() function (missMDA R in the BIOS pipeline). Next, we removed methylation outliers (range: 291-505) that exceeded three times the interquartile range (ewaff R) [45] in each dataset. We then combined the methylation β-values from all three imputed datasets and obtained residual methylation levels by regressing the effects of sex, age, percentages of epithelial and natural killer cells, EPIC array row, and bisulfite sample plate. In the combined dataset of imputed and residualized DNA methylation sites, we calculated the top 10% most variable methylation sites (72,889) to retain for further analysis (**Supplementary Figure 1, Supplementary Table 2**).

### Genotyping and calculation of polygenic scores

Genotyping was done according to the manufacturer’s protocols in 3,124 NTR samples on multiple genotyping platforms. Sample and SNP QC was performed per genotyping platform (build 37) for all individuals in the NTR. Genotype data were imputed to 1000 genomes phase 3 (v.5) [46] and a combined HRC 1.1 (EGA version) [47, 48] and GONL (v.4) [49] reference panel. The first was the basis for calculation of genetic principal components (PCs) and the second for calculation of PGSs. More details on QC and imputation are included in **Supplementary Note 1** and on calculation of genetic PCs in **Supplementary Note 2**.

Parental and offspring PGSs for schizophrenia [50], smoking initiation [51], educational attainment [52], social deprivation [53], BMI [54],and height [54] were calculated based on GWASs, that all omitted NTR from the discovery (meta-)analysis. Before generating the PGSs, QC steps involved removing SNPs with Hardy-Weinberg equilibrium (HWE) p-values below 0.00001, Mendelian error rates above 1%, genotype call rate below 98%, effect allele frequencies below 0.01 and above 0.99, imputation info below 0.10 and SNPs showing over 2% differences in allele frequencies between genotyping platforms. We retained 7,086,504 (schizophrenia), 6,982,922 (smoking initiation), 7,118,026 (EA), 6,997,217 (social deprivation), 2,220,360 (BMI), and 2,206,961 (height) SNPs.

LD-weighted betas were calculated from the processed summary statistics in the LDpred package (v.0.9) to correct for the effects of LD, and to maximize predictive accuracy of the PGS [55]. We randomly selected 2,500 individuals unrelated to the 2^nd^ degree from the NTR as a reference population to obtain LD patterns in an NTR genotype subset of well imputed variants. Weights to obtain the PGSs in PLINK2 (--score) [56] were calculated with an LD pruning window of 250 KB and the infinitesimal prior (LDpred-inf). We calculated the offspring, maternal, and paternal PGS for the 6 traits (18 PGSs total). Two dummy coded genotyping platforms and the first 10 genetic PCs were regressed on the PGSs. The standardized residuals (unit variance and zero means) were analyzed. If maternal or paternal polygenic scores were missing, we applied mean imputation (i.e., imputed the parental PGS with zero) [2] (*N_dataset1_* = 31; *N_dataset2_*= 213; *N_dataset3_* = 87).

### Analyses

As shown in Okbay et al. (2022) [9] (see also **Supplementary Note 3**), models that include the PGSs from offspring and two parents provide estimates of direct and indirect genetic effects, and allow for differences in maternal and paternal indirect genetic effects [9, 10].

Direct and indirect genetic effects for a single trait can be estimated by fitting the following regression model:

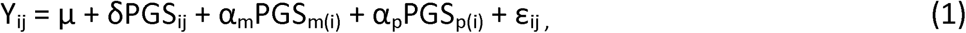

where Y_ij_ is the residualized methylation probe β-value, and i and j index family and individual; μ is the intercept, δ is the direct effect of the offspring PGS; α_m_ captures the maternal indirect genetic effects; α_p_ captures the paternal indirect genetic effects; PGS_m(i)_ and PGS_p(i)_ are the PGSs of the mother and father in family i; and ε_ij_ is a residual term.

We extended equation 1 by including maternal, paternal, and offspring PGS for multiple traits. This allows for the unbiased estimation of the direct and indirect genetic effects for a given complex trait, while accounting for direct and indirect effects on all other traits in the model.

The direct and indirect genetic effects for multiple traits are modelled as follows:

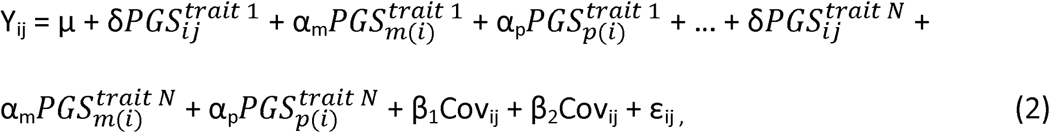

where subscript trait 1 through trait N indicates the trait for which the standardized residual polygenetic score was calculated and Cov_ij_ and Cov*_i_*_j_ indicate the DNA methylation dataset.

We tested the association between the 72,889 CpGs and the offspring, maternal, and paternal PGSs for the six traits simultaneously (equation 2), i.e., 18 predictors, using Generalized Estimation Equation (GEE) regression modeling to correct the standard errors for family clustering of individuals [57]. All analyses were carried out in R (version 4.2.2) [58] in the R package ‘gee’ (version 4.13-26) [59]. A Bonferroni correction was applied for the number of independent DNA methylation probes tested (*α* = 0.05/N independent variables), following the procedure outlined by Nyholt (2004) [60], where the number of independent tests is determined by Matrix Spectral Decomposition (MSD) of the imputed CpGs. The correlated CpGs could be reduced to 1,850 independent linear combinations of the probes and we set the significance level to (0.05/1,850) = 2.7 × 10^−5^. When analyzing the 72,889 CpGs, the regression model for 192 CpGs did not converge. We present the results for the remaining 72,697 successful analyses.

Significant CpGs from these analyses were followed up in four analyses. First, we queried the EWAS Atlas database to identify with which traits these CpGs had been previously associated [61]. Second, we queried the GoDMC database (http://mqtldb.godmc.org.uk/) for associated blood *cis* and *trans* methylation quantitative trait loci (mQTLs) for these CpGs [15] and examined if the identified mQTLs were mapped to genome-wide significant (*p* < 5 × 10^−8^) SNPs for the respective traits. Third, we investigated whether the DNA methylation levels in buccal cells and prefrontal cortex were significantly correlated (Spearman rank correlations at False Discovery Rate (FDR) *q* < 0.05) in a published dataset of matched post-mortem samples (*N* = 120) [62]. Fourth, we performed a look-up of the genes annotated to the significantly associated CpGs to investigate whether they are protein expressing in the human dorsal lateral prefrontal cortex and if so, whether these proteins are differentially regulated in schizophrenia [63]. Last, we performed trait enrichment analyses of the top 100 most significant sites identified in the mega-analysis for each PGS and complex trait in the EWAS Atlas.

## Results

Participant characteristics are described in **Table 1**. We analyzed data from 1,528 offspring and their parents (51.5% boys, mean [SD] age = 10 [2.8] years) to test the simultaneous association of offspring, maternal, and paternal PGSs for six complex traits with 72,697 buccal DNA methylation sites in offspring. The correlations between PGSs of different traits ranged from -0.29 and 0.23 (mean = -0.004, median = -0.01), with the strongest correlation observed between the Maternal PGSs of smoking initiation and EA (**Figure 1**). We accounted for these correlations by fitting the PGSs of all traits simultaneously in one model. The correlations between the offspring and parental PGSs for the same trait were approximately 0.5 as expected (range = 0.43-0.53, mean = 0.48, median = 0.47), while correlations between parental PGSs for the same trait were on average 0.04 (median = 0.04, range = -0.02-0.10).

**Figure 1.**
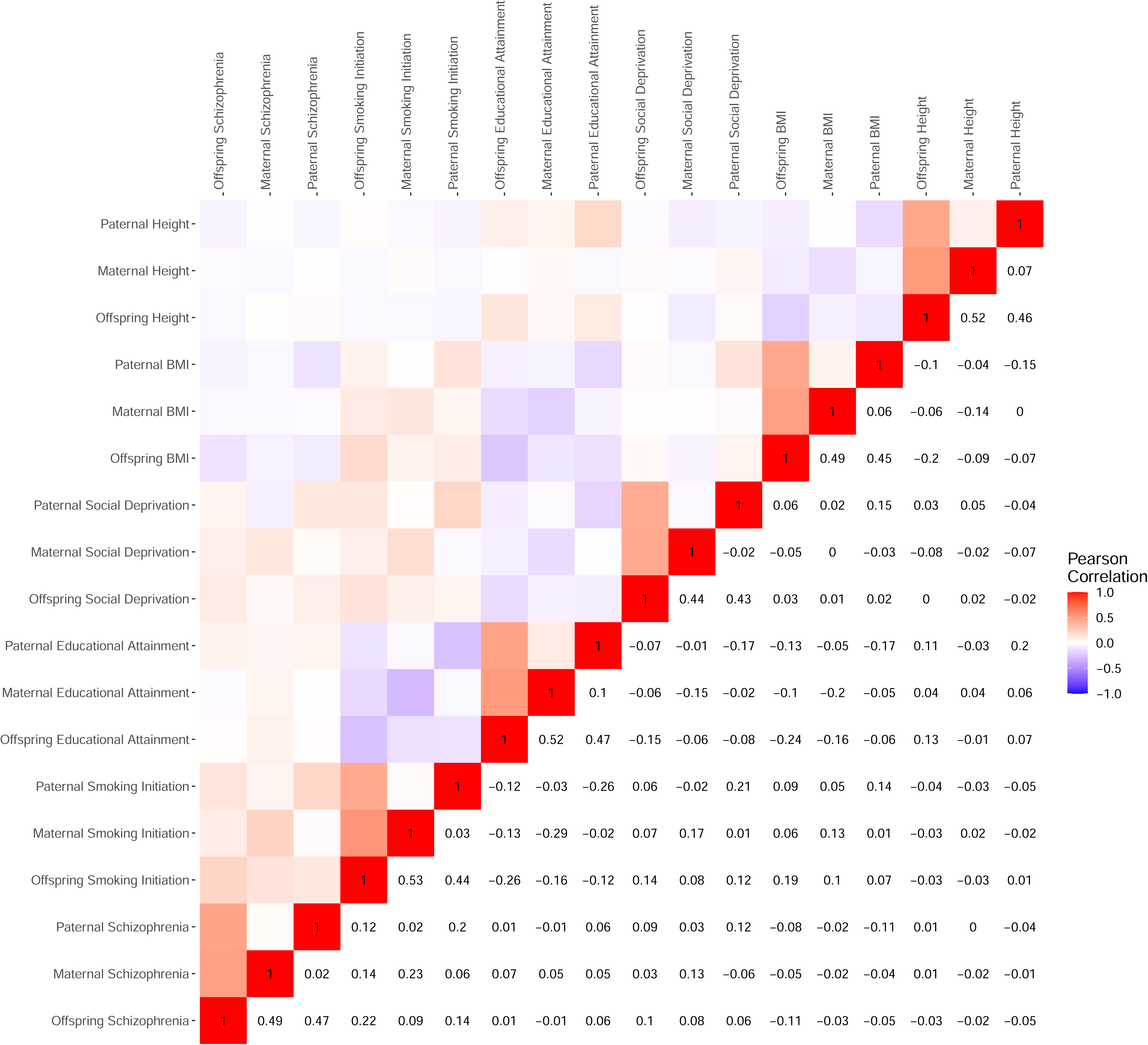
Pearson correlations among the offspring, maternal, and paternal polygenic scores for schizophrenia, smoking initiation, educational attainment, social deprivation, body mass index (BMI), and height.

**Table 1.**
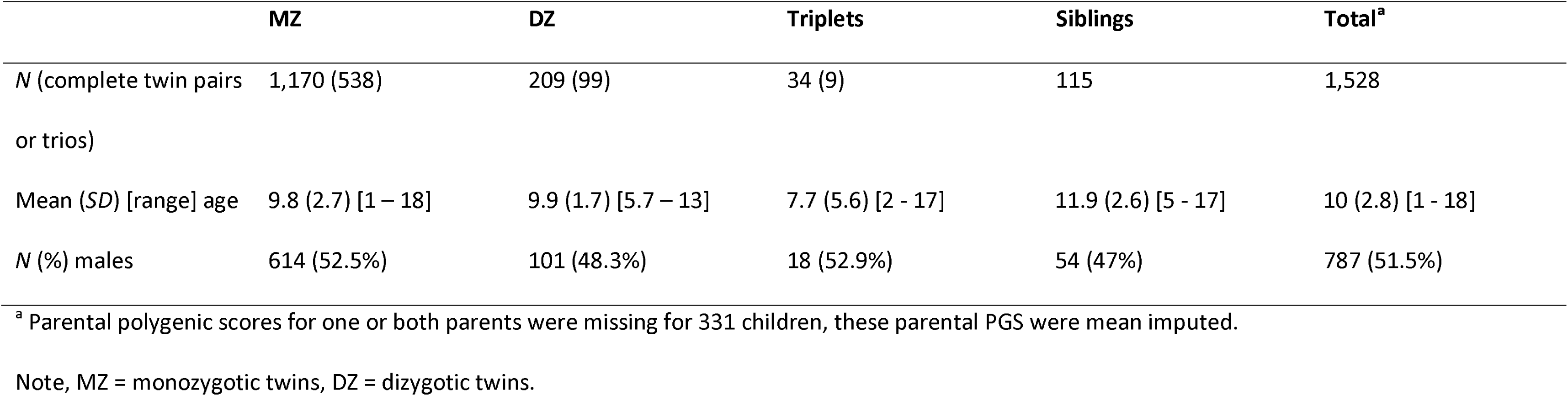
Demographics of the individuals included across the three buccal DNA methylation datasets in this study.

|  | **MZ** | **DZ** | **Triplets** | **Siblings** | **Total<sup>a</sup>** |
| --- | --- | --- | --- | --- | --- |
| *N* (complete twin pairs or trios) | 1,170 (538) | 209 (99) | 34 (9) | 115 | 1,528 |
| Mean (*SD*) [range] age | 9.8 (2.7) [1 – 18] | 9.9 (1.7) [5.7 – 13] | 7.7 (5.6) [2 - 17] | 11.9 (2.6) [5 - 17] | 10 (2.8) [1 - 18] |
| *N* (%) males | 614 (52.5%) | 101 (48.3%) | 18 (52.9%) | 54 (47%) | 787 (51.5%) |

<sup>a</sup> Parental polygenic scores for one or both parents were missing for 331 children, these parental PGS were mean imputed.
Note, MZ = monozygotic twins, DZ = dizygotic twins.

We observed independent significant associations between offspring methylation sites and maternal PGSs for schizophrenia (15), height (2), and BMI (1), and paternal PGSs for schizophrenia (1), EA (2), and height (4), (**Figure 2, Table 2**), demonstrating a mechanism for indirect effects from parental phenotype to the offspring methylome. The own offspring PGSs for schizophrenia, EA, and height showed associations with 21, 3, and 2 offspring methylation sites, respectively. None of the CpGs associated with maternal, paternal, or offspring PGSs overlapped. Thus, in total, 25 and 26 significant CpG sites (*p* < 2.7 × 10^−5^, Bonferroni correction for 1,850 tests) were associated with indirect and direct genetic effects across the six complex traits, respectively (**Supplementary Figures 2-7, Supplementary Table 3**).

**Figure 2.**
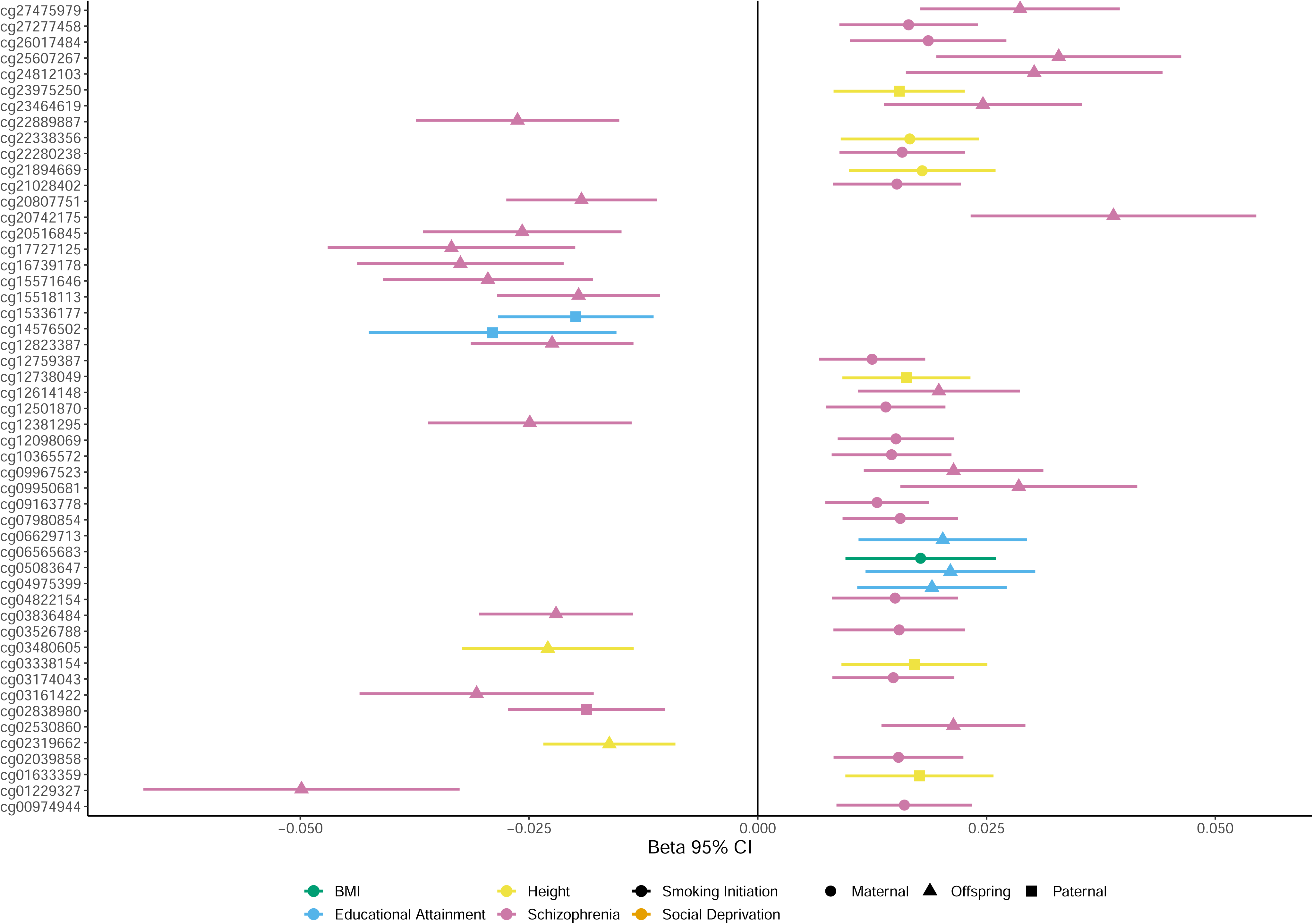
Buccal CpGs significantly associated (alpha = 2.7 × 10^−5^) with maternal (⍰), paternal (⍰), or offspring (Δ) polygenic scores for schizophrenia, educational attainment, body mass index (BMI), and height.

**Table 2.** Buccal CpGs significantly associated (alpha = 2.7 × 10^−5^) with maternal, paternal, or offspring polygenic scores for schizophrenia, educational attainment (EA), body mass index (BMI), and height.

| PGS | CpG | CHR:BP<sup>a</sup> | Gene<sup>b</sup> | mQTLs | SNPs | Complex traits |
| --- | --- | --- | --- | --- | --- | --- |
| Maternal Schizophrenia | cg00974944 | 3:61649924 | *PTPRG* |  |  |  |
| Maternal Schizophrenia | cg02039858 | 14:65828769 |  |  |  |  |
| Maternal Schizophrenia | cg03174043 | 2:20548412 |  | 185 |  | fractional exhaled nitric oxide |
| Maternal Schizophrenia | cg03526788 | 8:130899478 | *FAM49B* |  |  |  |
| Maternal Schizophrenia | cg04822154 | 8:124692680 |  |  |  |  |
| Maternal Schizophrenia | cg07980854 | 20:57586338 |  | 634 |  | breast cancer; type 2 diabetes; atopy; psoriasis |
| Maternal Schizophrenia | cg09163778 | 6:31125417 | *TCF19; CCHCR1* |  |  |  |
| Maternal Schizophrenia | cg10365572 | 5:141245930 | *PCDH1* |  |  |  |
| Maternal Schizophrenia | cg12098069 | 13:42537172 | *VWA8-AS1* |  |  |  |
| Maternal Schizophrenia | cg12501870 | 8:81803721 |  | 182 |  |  |
| Maternal Schizophrenia | cg12759387 | 1:27849177 |  | 177 |  | systemic lupus erythematosus; polychlorinated biphenyls exposure |
| Maternal Schizophrenia | cg21028402 | 6:113932902 |  | 297 |  | helicobacter pylori infection; oral squamous cell carcinoma |
| Maternal Schizophrenia | cg22280238 | 17:79251323 | *SLC38A10* |  |  |  |
| Maternal Schizophrenia | cg26017484 | 1:3322309 | *PRDM16* |  |  | adenoma |
| Maternal Schizophrenia | cg27277458 | 10:63597194 |  |  |  |  |
| Paternal Schizophrenia | cg02838980 | 16:10402487 |  |  |  |  |
| Offspring Schizophrenia | cg01229327 | 19:54311998 | *NLRP12* |  |  |  |
| Offspring Schizophrenia | cg02530860 | 8:144371537 |  | 646 |  | gender; sperm viability; assisted reproduction technology; infant sex; sexually dimorphic |
| Offspring Schizophrenia | cg03161422 | 4:175256045 |  |  |  |  |
| Offspring Schizophrenia | cg03836484 | 10:25349182 | *ENKUR* |  |  |  |
| Offspring Schizophrenia | cg09950681 | 11:393392 | *PKP3* |  |  |  |
| Offspring Schizophrenia | cg09967523 | 18:18943072 | *GREB1L* |  |  |  |
| Offspring Schizophrenia | cg12381295 | 20:3880297 | *PANK2* |  |  | gingivo-buccal oral squamous cell carcinoma (OSCC-GB) |
| Offspring Schizophrenia | cg12614148 | 1:118325026 |  | 98 |  |  |
| Offspring Schizophrenia | cg12823387 | 1:214614656 | *PTPN14* |  |  |  |
| Offspring Schizophrenia | cg15518113 | 1:167400121 | *CD247* | 105 |  | Air pollution (NO<sub>2</sub>); adrenocortical carcinoma; smoking; multiple sclerosis; |
| Offspring Schizophrenia | cg15571646 | 2:238767351 | *RAMP1* |  |  |  |
| Offspring Schizophrenia | cg16739178 | 16:85470674 |  | 50 |  | papillary thyroid carcinoma; aging; BMI; Crohn's disease; Behcet's disease; maternal hypertensive disorders in pregnancy; gestational age; blood C-reactive protein level; inflammatory bowel disease |
| Offspring Schizophrenia | cg17727125 | 8:38441156 |  |  |  |  |
| Offspring Schizophrenia | cg20516845 | 21:43823604 | *UBASH3A* | 140 |  |  |
| Offspring Schizophrenia | cg20742175 | 20:57267357 | *NPEPL1*;<br>*STX16-NPEPL1* |  |  | high risk cutaneous squamous cell carcinoma (cSCC) |
| Offspring Schizophrenia | cg20807751 | 9:84025287 |  |  |  |  |
| Offspring Schizophrenia | cg22889887 | 11:113433931 |  | 6 |  |  |
| Offspring Schizophrenia | cg23464619 | 13:95513506 |  | 50 |  | hepatocellular carcinoma; colorectal laterally spreading tumor; inflammatory bowel disease; ulcerative colitis |
| Offspring Schizophrenia | cg24812103 | 8:101735630 | *PABPC1* | 244 |  | prenatal arsenic exposure; gestational diabetes mellitus; inflammatory bowel disease |
| Offspring Schizophrenia | cg25607267 | 12:122362708 | *WDR66* |  |  |  |
| Offspring Schizophrenia | cg27475979 | 16:89471039 | *ANKRD11* |  |  |  |
| Paternal EA | cg14576502 | 8:102962279 | *NCALD* |  |  | recurrent adult-type IDH-mutant gliomas |
| Paternal EA | cg15336177 | 1:99369228 | *LPPR5* |  |  |  |
| Offspring EA | cg04975399 | 1:67822225 | *IL12RB2* |  |  | fractional exhaled nitric oxide |
| Offspring EA | cg05083647 | 1:204897364 | *NFASC* |  |  |  |
| Offspring EA | cg06629713 | 2:60580559 |  | 43 |  | primary Sjogren's Syndrome; obesity; Type 2 diabetes; asthma; atopy; Crohn's disease; fractional exhaled nitric oxide; hepatitis B virus-related liver disease progression |
| Maternal BMI | cg06565683 | 20:46817989 |  |  |  |  |
| Maternal Height | cg21894669 | 2:165379442 | *GRB14* |  |  |  |
| Maternal Height | cg22338356 | 8:48739161 | *PRKDC* |  |  | Down Syndrome; Fetal alcohol spectrum disorder |
| Paternal Height | cg01633359 | 5:33439359 |  | 164 | 56 |  |
| Paternal Height | cg03338154 | 6:41121714 | TREML1 |  |  |  |
| Paternal Height | cg12738049 | 15:76509692 | ETFA |  |  |  |
| Paternal Height | cg23975250 | 22:44757380 |  |  |  | sexually dimorphic |
| Offspring Height | cg02319662 | 4:108127694 |  |  |  |  |
| Offspring Height | cg03480605 | 7:151038446 | NUB1 | 216 | 17 | maternal smoking; obesity; oral squamous cell carcinoma |

<sup>a</sup>CHR:BP = Chromosome base pair location in genome build 37. <sup>b</sup>Gene = NCBI gene summaries and Ensembl links are added to **Supplementary Table 8**. mQTLs = number of blood *cis* methylation quantitative trait loci (mQTLs) [15] associated with the CpGs. SNPs = number of genome-wide significant ( $p < 5 \times 10^{-8}$ ) SNPs overlapping with blood *cis* mQTLs for the CpGs. Complex traits = complex traits previously associated with the CpGs in the EWAS Atlas [61].

The query of the EWAS Atlas database [61] for the 37 CpGs associated with direct or indirect genetic effects for schizophrenia showed that 12 CpGs (5 associated with the maternal and 7 with the offspring PGS) had known trait associations, though not with schizophrenia or other mental health traits (**Table 2**). Similarly, 3 out of the 5 CpGs associated with paternal (2) or offspring (3) PGSs for EA and 3 out of the 8 CpGs associated with maternal (2), paternal (4), or offspring (2) PGSs for height were included in the EWAS Atlas but showed no previous associations with the respective or a related complex trait. Trait enrichment analyses for the top 100 CpGs associated with offspring and parental PGSs for all six complex traits showed enrichment for 5 to 18 traits per PGS (mean = 10.2, median = 10) with on average 2.2 CpGs associated per trait (range = 1-7, median = 2, **Supplementary Table 4**). Notably, many of the enriched traits are phenotypically associated with the relevant PGSs, such as enrichment for bipolar and major depressive disorder and maternal hypertensive disorders in pregnancy for CpGs associated with maternal indirect genetic effects for schizophrenia and household socioeconomic status in childhood for CpGs with direct and indirect genetic effects for schizophrenia.

A query of the GoDMC blood mQTL database [15] with the 51 significant CpGs identified 3,237 *cis* mQTLs for 16 CpGs (mean = 202.3 mQTLs per associated CpG, median = 170.5) associated with schizophrenia (13 CpGs), EA (1), and height (2), with roughly half (1,639/3,237) of the mQTLs associated with indirect genetic effect CpGs (**Table 2, Supplementary Table 5**). Of the 164 mQTLs associated with cg01633359 (paternal PGS for height) and 216 of the mQTLs associated with cg03480605 (offspring PGS for height) 56 and 17, respectively, had previously been associated with height at genome-wide significant levels (*p* < 5 × 10^−8^, **Table 2, Supplementary Figure 8, Supplementary Table 6**) [54]. None of the mQTLs for the schizophrenia and EA associated CpGs were previously found to be significantly associated with their respective traits at genome-wide significant levels [50, 52].

We looked at CpGs significantly associated with direct or indirect genetic effects in a published dataset (*N* = 120) of correlations between DNA methylation levels in buccal cells and the prefrontal cortex [62]. One CpG associated with the maternal PGS for schizophrenia (cg09163778, *r* = 0.31, *p* = 5.86 × 10^−4^), 2 CPGs associated with the offspring PGS for schizophrenia (cg02530860, *r* = 0.67, *p* = 7.51 × 10^−17^, cg01229327, *r* = 0.49, *p* = 1.91 × 10^−8^), and one CpG associated with the paternal PGS for height (cg01633359, *r* = 0.29, *p* = 1.44 × 10^−3^) showed significantly (FDR *q* < 0.05) correlated DNA methylation levels between buccal cells and the prefrontal cortex (**Supplementary Table 7**). Because of the relatively large number of CpGs associated with schizophrenia PGSs, we performed further follow-up analyses on the differential regulation in schizophrenia of the proteins encoded by the 29 genes annotated to these CpGs. Of these genes, 9 genes had protein expression detected in the dorsal lateral prefrontal cortex (dlPFC) and 2 of those (*PRKDC* and *NCALD*) were significantly differentially regulated in schizophrenia (**Supplementary Table 8**). Neither gene was annotated to a CpG associated with the offspring or parental PGS for schizophrenia, rather *PRKDC* was annotated to cg22338356 (chr8:48739161) which was associated with the maternal PGS for height, and *NCALD* was annotated to cg14576502 (chr8:102962279) which was associated with the paternal PGS for EA.

## Discussion

We investigated direct and indirect genetic influences on offspring DNA methylation for schizophrenia, smoking initiation, EA, social deprivation, BMI, and height in a cohort of 1,528 children and their parents. We identified 37 CpGs significantly associated with schizophrenia, 5 with EA, 1 with BMI, and 8 with height. None of the identified CpGs overlapped between polygenic scores. We found the strongest genetic nurture effect for schizophrenia. Of the schizophrenia associated CpGs, 43% (16 CpGs, 93.7% maternal) showed an association with parental PGSs. Overall, the maternal genetic nurture effect for schizophrenia (15 CpGs) on offspring DNA methylation represented 83.3% of all maternal genetic nurture effects (18 CpGs) and 58% of all genetic nurture effects (25 CpGs).

We identified 25 CpGs associated with parental PGSs. These results indicate that one mechanism for genetic nurture is via the offspring methylome, though the relation of the identified CpGs with offspring outcomes remains to be determined. Of these, 18 CpGs were specific to associations with maternal PGSs, and 7 were specific to paternal PGSs, suggesting that genetically driven paternal behaviors have a smaller impact on offspring DNA methylation. The larger number of CpGs associated with maternal PGSs for schizophrenia (15/16 CpGs) is consistent with a stronger influence of genetic nurture effects on offspring DNA methylation in utero. This aligns with the “Developmental Origins of Health and Disease” (DOHad) hypothesis, that prenatal environmental factors have long-term effects through epigenetic mechanisms [64]. Unfavorable intrauterine conditions leading to poor fetal growth are particularly relevant to the DOHaD hypothesis [65, 66].

Observational studies indicate that the intrauterine environment is influenced by maternal height and weight, and both direct and maternal indirect genetic effects affect offspring birth length, weight, and gestational age [67, 68]. The CpGs associated with maternal PGSs for height (2 CpGs) and BMI (1 CpG) may be involved in these processes. However, recent research suggests that the impact of intrauterine growth restriction on complex disease risk is limited, and that non-transmitted maternal genetic factors for birth weight do not contribute to offspring disease risk beyond genetic pleiotropy with offspring genetic factors [69]. This aligns with our finding that the largest number of findings for non-transmitted maternal genetic factors is not for body size but for schizophrenia. Most CpGs associated with indirect genetic effects were observed for maternal PGSs, particularly for schizophrenia. This observation may result from both pre-and postnatal exposures, including rearing environment and maternal parenting behaviors. One direction for further research is to examine DNA methylation in neonatal samples to better isolate prenatal from postnatal effects.

Several of the CpG sites associated with the offspring and parental PGSs for schizophrenia annotate to genes implicated in neurodevelopmental and immune processes (**Table 2, Supplementary Table 8**), consistent with the pathophysiology of schizophrenia [70, 71]. For instance, *PTPRG* (cg00974944) and *PCDH1* (cg10365572), both associated with the maternal schizophrenia PGS, are involved in cell growth regulation and neural adhesion, respectively, supporting the role of dysregulated synaptic processes in schizophrenia risk [50]. Similarly, *CCHCR1* (cg09163778, associated with maternal schizophrenia PGS), located within the major histocompatibility complex (MHC) region, is of particular interest due to the MHC’s established role in immune regulation and its prior associations with schizophrenia [72].

For EA, several genes associated with CpG sites suggest mechanisms related to neuronal signaling and synaptic plasticity (**Table 2, Supplementary Table 8**). For example, *NCALD* (cg14576502, associated with the paternal PGS for EA), a gene encoding a calcium-binding protein involved in G-protein-coupled receptor signal transduction, may influence cognitive functions through its regulatory role in calcium signaling, a key process in synaptic activity and plasticity [73]. We also observed significant regulation of *NCALD* protein expression in the dlPFC in schizophrenia.

For height, several CpG sites annotate to genes related to growth, metabolism, and cellular signaling (**Table 2, Supplementary Table 8**). *GRB14* (cg21894669, associated with maternal PGS for height), encodes a growth factor receptor-binding protein that regulates insulin receptor signaling. By modulating receptor tyrosine kinase activity, particularly in insulin pathways, it likely influences growth and metabolism. Additionally, *PRKDC* (cg22338356, associated with the maternal PGS for height), one of the two genes with significantly regulated protein expression in the dlPFC in schizophrenia, is highly expressed in immune cells and mutations have been linked to autoimmune diseases and increased immune response [74]. This connection between immune system regulation, schizophrenia, and height reflects previous findings from GWAS studies [75].

We did not model a relation of the identified CpGs with offspring outcomes, but explored the literature to determine whether the significant CpGs for each trait had been associated with the PGS for that trait and found that none of the CpGs significantly associated with direct or indirect genetic effects had been associated with polygenic scores for schizophrenia or BMI [25, 29–31]. These previous studies all comprised adults, whereas our study focused on children. The previous studies on schizophrenia mainly comprised adult patient populations focused on DNA methylation in blood or post-mortem brain samples (*N* range = 88-847). DNA methylation is often tissue specific and despite identifying blood *cis* mQTLs for approximately one-third of the CpGs associated with direct and indirect genetic effects, some *cis* mQTLs are not found in blood. Aside from tissue-specificity, the lack over overlap between CpGs associated in buccal and blood *cis* mQTLs might reflect differences in DNA methylation arrays. Previously, we found that 46.3% of buccal mQTLs as measured on the Illumina EPIC array overlap with blood mQTLs as measured on the Illumina 450K array when 15,897 of 33,749 EPIC sites were present on the 450K [36]. As blood and buccal *cis* mQTLs have a high (71%) concordance, we expect to have identified most *cis* mQTLs [76]. None of the identified CpGs were associated with *trans* mQTLs. Three CpGs associated with schizophrenia PGSs (1 maternal, 2 offspring) and one CpG associated with paternal PGS for height showed correlated methylation levels between buccal cells and prefrontal cortex brain samples [62]. Future studies should investigate direct and indirect genetic effects in multiple tissues relevant to health outcomes to better understand tissue-specific versus tissue-agnostic DNA methylation signatures.

This study is the first to test if indirect genetic effects influence DNA methylation in offspring. It also explored the direct genetic effects on DNA methylation in children. We analyzed data from 1,528 children and their parents. Most children were related to at least one other person in the sample, such as their co-twin and thus the effective sample size is smaller than 1,528. Given the sample size, we took three measures to reduce the multiple testing burden. First, we focused on the top 10% most variable CpGs, which have been shown to be the most reliable [32]. Second, we adjusted our multiple testing threshold based on the number of independent CpGs, considering the high correlations between them. Third, our approach of modeling multiple complex traits simultaneously allowed us to avoid correcting for the number of traits investigated, which would have been necessary in a separate modeling strategy. By limiting our investigation to highly variable CpGs, we cannot draw conclusions on less variable CpGs or CpGs with strong temporal patterns. Overall, our application highlights its potential to investigate epigenetic mechanisms of indirect genetic effects and encourages testing in large family cohorts, across different tissues, and populations.

In summary, we found support for the hypothesis that indirect genetic effects are associated with DNA methylation in offspring. By simultaneously modelling multiple complex traits, we found robust associations of indirect genetic effects on offspring DNA methylation for schizophrenia, EA, BMI, and height, but not smoking initiation and social deprivation. Most of these associations were discovered in relation to maternal PGSs, consistent with the interpretation that the prenatal environment influences the DNA methylation of offspring during embryonic development or reflect a stronger maternal influence on the offspring’s environment during the pre- and perinatal period and early childhood.

## Data Availability

The standardized protocol for large-scale collection of buccal-cell (and urine) samples in the home situation as developed for the ACTION Biomarker Study in children is available at https://dx.doi.org/10.17504/protocols.io.eq2ly7qkwlx9/v1. The data of the Netherlands Twin Register (NTR) may be accessed, upon approval of the data access committee, through the NTR (https://ntr-data-request.psy.vu.nl/).

## Declarations

## Acknowledgments

The Netherlands Twin Register (NTR) warmly thanks all twin families for their participation and are grateful to all researchers involved in the data collection for the ACTION Biomarker Study. Additionally, we thank Cato Romero, MSc. for feedback during the revision of this manuscript. The current work is supported by the Consortium on Individual Development (CID) and the “Aggression in Children: Unraveling gene-environment interplay to inform Treatment and InterventiON strategies” project (ACTION). CID is funded through the Gravitation Program of the Dutch Ministry of Education, Culture, and Science and the Netherlands Organization for Scientific Research (NWO grant number 024-001-003). ACTION received funding from the European Union Seventh Framework Program (FP7/2007-2013) under grant agreement no 602768. M.B. is supported by an ERC consolidator grant (WELL-BEING 771057, PI Bartels) and NWO VICI grant (VI.C.2111.054, PI Bartels). J.D. acknowledges the NWO-funded X-omics project (184.034.019) and D.I.B. the Royal Netherlands Academy of Arts and Sciences Professor Award (PAH/6635).

## Authors’ contributions

Conceptualization, F.A.H., R.P., J.J.H., C.V.D., J.D. and D.I.B.; methodology, C.V.D; formal analysis, F.A.H., R.P. and A.B.S.; investigation, F.A.H., M.B. and D.I.B.; resources, R.P., A.J.V.A., E.A.E., J.J.H. and J.D.; writing—original draft preparation, F.A.H., R.P., J.J.H., C.V.D., J.D. and D.I.B.; writing—review and editing, F.A.H., R.P., A.J.V.A., A.B.S., M.B., J.J.H., C.V.D., J.D. and D.I.B.; visualization, F.A.H. and C.V.D.; supervision, J.D. and D.I.B.; funding acquisition, M.B. and D.I.B. All authors have read and agreed to the published version of the manuscript.

## Availability of data and code

The standardized protocol for large scale collection of buccal-cell (and urine) samples in the home situation as developed for the ACTION Biomarker Study in children is available at https://dx.doi.org/10.17504/protocols.io.eq2ly7qkwlx9/v1. The data of the Netherlands Twin Register (NTR) may be accessed, upon approval of the data access committee, through the NTR (https://ntr-data-request.psy.vu.nl/). The pipeline for DNA methylation–array analysis developed by the Biobank-based Integrative Omics Study (BIOS) consortium is available at https://molepi.github.io/DNAmArray_workflow/ (https://doi.org/10.5281/zenodo.3355292). All software used to perform the analyses are available online. All scripts used to run the analyses (empirical and simulated) are available at our GitHub (https://github.com/FionaAHagenbeek/MethylationPGS).

## References

1. Bates TC, Maher BS, Medland SE, McAloney K, Wright MJ, Hansell NK, et al. The Nature of Nurture: Using a Virtual-Parent Design to Test Parenting Effects on Children’s Educational Attainment in Genotyped Families. Twin Research and Human Genetics. 2018;21:73–83.

2. Kong A, Thorleifsson G, Frigge ML, Vilhjalmsson BJ, Young AI, Thorgeirsson TE, et al. The nature of nurture: Effects of parental genotypes. Science. 2018;359:424–428.

3. Balbona JV, Kim Y, Keller MC. Estimation of Parental Effects Using Polygenic Scores. Behav Genet. 2021;51:264–278.

4. Cavalli-Sforza LL, Feldman MW. Cultural transmission and evolution: a quantitative approach. Monogr Popul Biol. 1981;16:1–388.

5. Eaves LJ. A model for sibling effects in man. Heredity. 1976;36:205–214.

6. Odling-Smee J, Laland KN. Ecological Inheritance and Cultural Inheritance: What Are They and How Do They Differ? Biol Theory. 2011;6:220–230.

7. McAdams TA, Cheesman R, Ahmadzadeh YI. Annual Research Review: Towards a deeper understanding of nature and nurture: combining family-based quasi-experimental methods with genomic data. Journal of Child Psychology and Psychiatry. 2023;64:693–707.

8. Swagerman SC, van Bergen E, Dolan C, de Geus EJC, Koenis MMG, Hulshoff Pol HE, et al. Genetic transmission of reading ability. Brain and Language. 2017;172:3–8.

9. Okbay A, Wu Y, Wang N, Jayashankar H, Bennett M, Nehzati SM, et al. Polygenic prediction of educational attainment within and between families from genome-wide association analyses in 3 million individuals. Nat Genet. 2022;54:437–449.

10. Tubbs JD, Porsch RM, Cherny SS, Sham PC. The Genes We Inherit and Those We Don’t: Maternal Genetic Nurture and Child BMI Trajectories. Behav Genet. 2020;50:310–319.

11. Bruins S, Dolan CV, Boomsma DI. The Power to Detect Cultural Transmission in the Nuclear Twin Family Design With and Without Polygenic Risk Scores and in the Transmitted– Nontransmitted (Alleles) Design. Twin Res Hum Genet. 2020;23:265–270.

12. Wang B, Baldwin JR, Schoeler T, Cheesman R, Barkhuizen W, Dudbridge F, et al. Robust genetic nurture effects on education: A systematic review and meta-analysis based on 38,654 families across 8 cohorts. The American Journal of Human Genetics. 2021;108:1780–1791.

13. Buescher JM, Driggers EM. Integration of omics: more than the sum of its parts. Cancer & Metabolism. 2016;4:4.

14. Carlberg C, Molnár F. What Is Epigenomics? In: Carlberg C, Molnár F, editors. Human Epigenomics, Singapore: Springer; 2018. p. 3–18.

15. Min JL, Hemani G, Hannon E, Dekkers KF, Castillo-Fernandez J, Luijk R, et al. Genomic and phenotypic insights from an atlas of genetic effects on DNA methylation. Nat Genet. 2021;53:1311–1321.

16. van Dongen J, Nivard MG, Willemsen G, Hottenga J-J, Helmer Q, Dolan CV, et al. Genetic and environmental influences interact with age and sex in shaping the human methylome. Nat Commun. 2016;7:11115.

17. Zhou FC, Resendiz M, Lo C-L. Chapter 31 - Environmental Influence of Epigenetics. In: Tollefsbol TO, editor. Handbook of Epigenetics (Second Edition), Academic Press; 2017. p. 477–494.

18. Weaver ICG, Cervoni N, Champagne FA, D’Alessio AC, Sharma S, Seckl JR, et al. Epigenetic programming by maternal behavior. Nat Neurosci. 2004;7:847–854.

19. Cecil CAM, Zhang Y, Nolte T. Childhood maltreatment and DNA methylation: A systematic review. Neuroscience & Biobehavioral Reviews. 2020;112:392–409.

20. Parade SH, Huffhines L, Daniels TE, Stroud LR, Nugent NR, Tyrka AR. A systematic review of childhood maltreatment and DNA methylation: candidate gene and epigenome-wide approaches. Transl Psychiatry. 2021;11:1–33.

21. Rubens M, Bruenig D, Adams JAM, Suresh SM, Sathyanarayanan A, Haslam D, et al. Childhood maltreatment and DNA methylation: A systematic review. Neuroscience & Biobehavioral Reviews. 2023;147:105079.

22. Christiansen C, Castillo-Fernandez JE, Domingo-Relloso A, Zhao W, El-Sayed Moustafa JS, Tsai P-C, et al. Novel DNA methylation signatures of tobacco smoking with trans-ethnic effects. Clinical Epigenetics. 2021;13:36.

23. van Dongen J, Willemsen G, BIOS Consortium, de Geus EJ, Boomsma DI, Neale MC. Effects of smoking on genome-wide DNA methylation profiles: A study of discordant and concordant monozygotic twin pairs. eLife. 2023;12:e83286.

24. Szyf M, McGowan P, Meaney MJ. The social environment and the epigenome. Environmental and Molecular Mutagenesis. 2008;49:46–60.

25. Hannon E, Dempster E, Viana J, Burrage J, Smith AR, Macdonald R, et al. An integrated genetic-epigenetic analysis of schizophrenia: evidence for co-localization of genetic associations and differential DNA methylation. Genome Biology. 2016;17:176.

26. Montano C, Taub MA, Jaffe A, Briem E, Feinberg JI, Trygvadottir R, et al. Association of DNA Methylation Differences With Schizophrenia in an Epigenome-Wide Association Study. JAMA Psychiatry. 2016;73:506–514.

27. Dick KJ, Nelson CP, Tsaprouni L, Sandling JK, Aïssi D, Wahl S, et al. DNA methylation and body-mass index: a genome-wide analysis. The Lancet. 2014;383:1990–1998.

28. Vehmeijer FOL, Küpers LK, Sharp GC, Salas LA, Lent S, Jima DD, et al. DNA methylation and body mass index from birth to adolescence: meta-analyses of epigenome-wide association studies. Genome Medicine. 2020;12:105.

29. Kiltschewskij DJ, Reay WR, Geaghan MP, Atkins JR, Xavier A, Zhang X, et al. Alteration of DNA Methylation and Epigenetic Scores Associated With Features of Schizophrenia and Common Variant Genetic Risk. Biological Psychiatry. 2023. 2023. 10.1016/j.biopsych.2023.07.010.

30. Viana J, Hannon E, Dempster E, Pidsley R, Macdonald R, Knox O, et al. Schizophrenia-associated methylomic variation: molecular signatures of disease and polygenic risk burden across multiple brain regions. Human Molecular Genetics. 2017;26:210–225.

31. Yu C, Hodge AM, Wong EM, Joo JE, Makalic E, Schmidt DF, et al. Does genetic predisposition modify the effect of lifestyle-related factors on DNA methylation? Epigenetics. 2022;17:1838–1847.

32. Van Asselt AJ, Beck JJ, Finnicum CT, Johnson BN, Kallsen N, Hottenga JJ, et al. Genome-Wide DNA Methylation Profiles in Whole-Blood and Buccal Samples—Cross-Sectional, Longitudinal, and across Platforms. International Journal of Molecular Sciences. 2023;24:14640.

33. Ligthart L, Beijsterveldt CEM van, Kevenaar ST, Zeeuw E de, Bergen E van, Bruins S, et al. The Netherlands Twin Register: Longitudinal Research Based on Twin and Twin-Family Designs. Twin Research and Human Genetics. 2019;22:623–636.

34. Bartels M, Hendriks A, Mauri M, Krapohl E, Whipp A, Bolhuis K, et al. Childhood aggression and the co-occurrence of behavioural and emotional problems: results across ages 3–16 years from multiple raters in six cohorts in the EU-ACTION project. Eur Child Adolesc Psychiatry. 2018;27:1105–1121.

35. Hagenbeek FA, Roetman PJ, Pool R, Kluft C, Harms AC, van Dongen J, et al. Urinary Amine and Organic Acid Metabolites Evaluated as Markers for Childhood Aggression: The ACTION Biomarker Study. Frontiers in Psychiatry. 2020;11.

36. van Dongen J, Ehli EA, Jansen R, van Beijsterveldt CEM, Willemsen G, Hottenga JJ, et al. Genome-wide analysis of DNA methylation in buccal cells: a study of monozygotic twins and mQTLs. Epigenetics & Chromatin. 2018;11:54.

37. Hagenbeek FA, Dongen J van, Roetman PJ, Ehli EA, Bartels M, Vermeiren RRJM, et al. ACTION Biomarker Study. ProtocolsIo. 2023. March 2023. 10.17504/protocols.io.eq2ly7qkwlx9/v1.

38. Moran S, Arribas C, Esteller M. Validation of a DNA methylation microarray for 850,000 CpG sites of the human genome enriched in enhancer sequences. Epigenomics. 2016;8:389–399.

39. van Dongen J, Hagenbeek FA, Suderman M, Roetman PJ, Sugden K, Chiocchetti AG, et al. DNA methylation signatures of aggression and closely related constructs: A meta-analysis of epigenome-wide studies across the lifespan. Mol Psychiatry. 2021;26:2148–2162.

40. Sinke L, van Iterson M, Cats D, Slieker R, Heijmans B. DNAmArray: Streamlined workflow for the quality control, normalization, and analysis of Illumina methylation array data. 2019.

41. van Iterson M, Tobi EW, Slieker RC, den Hollander W, Luijk R, Slagboom PE, et al. MethylAid: visual and interactive quality control of large Illumina 450k datasets. Bioinformatics. 2014;30:3435–3437.

42. van Iterson M, Cats D, Hop P, BIOS Consortium, Heijmans BT. omicsPrint: detection of data linkage errors in multiple omics studies. Bioinformatics. 2018;34:2142–2143.

43. Min JL, Hemani G, Davey Smith G, Relton C, Suderman M. Meffil: efficient normalization and analysis of very large DNA methylation datasets. Bioinformatics. 2018;34:3983–3989.

44. Zheng SC, Webster AP, Dong D, Feber A, Graham DG, Sullivan R, et al. A novel cell-type deconvolution algorithm reveals substantial contamination by immune cells in saliva, buccal and cervix. Epigenomics. 2018;10:925–940.

45. Suderman, Matthew, Sharp, Gemma, Yousefi, Paul, Kupers, Leanne. perishky/ewaff: Efficient and Flexible EWAS version 0.0.2 from GitHub. 2019.

46. Auton A, Abecasis GR, Altshuler DM, Durbin RM, Abecasis GR, Bentley DR, et al. A global reference for human genetic variation. Nature. 2015;526:68–74.

47. McCarthy S, Das S, Kretzschmar W, Delaneau O, Wood AR, Teumer A, et al. A reference panel of 64,976 haplotypes for genotype imputation. Nat Genet. 2016;48:1279–1283.

48. The Haplotype Reference Consortium - EGA European Genome-Phenome Archive. https://ega-archive.org/studies/EGAS00001001710. Accessed 6 April 2023.

49. Boomsma DI, Wijmenga C, Slagboom EP, Swertz MA, Karssen LC, Abdellaoui A, et al. The Genome of the Netherlands: design, and project goals. Eur J Hum Genet. 2014;22:221–227.

50. Trubetskoy V, Pardiñas AF, Qi T, Panagiotaropoulou G, Awasthi S, Bigdeli TB, et al. Mapping genomic loci implicates genes and synaptic biology in schizophrenia. Nature. 2022;604:502–508.

51. Liu M, Jiang Y, Wedow R, Li Y, Brazel DM, Chen F, et al. Association studies of up to 1.2 million individuals yield new insights into the genetic etiology of tobacco and alcohol use. Nat Genet. 2019;51:237–244.

52. Lee JJ, Wedow R, Okbay A, Kong E, Maghzian O, Zacher M, et al. Gene discovery and polygenic prediction from a genome-wide association study of educational attainment in 1.1 million individuals. Nat Genet. 2018;50:1112–1121.

53. Hill WD, Hagenaars SP, Marioni RE, Harris SE, Liewald DCM, Davies G, et al. Molecular Genetic Contributions to Social Deprivation and Household Income in UK Biobank. Current Biology. 2016;26:3083–3089.

54. Yengo L, Sidorenko J, Kemper KE, Zheng Z, Wood AR, Weedon MN, et al. Meta-analysis of genome-wide association studies for height and body mass index in ∼700000 individuals of European ancestry. Hum Mol Genet. 2018;27:3641–3649.

55. Vilhjálmsson BJ, Yang J, Finucane HK, Gusev A, Lindström S, Ripke S, et al. Modeling Linkage Disequilibrium Increases Accuracy of Polygenic Risk Scores. The American Journal of Human Genetics. 2015;97:576–592.

56. Chang CC, Chow CC, Tellier LC, Vattikuti S, Purcell SM, Lee JJ. Second-generation PLINK: rising to the challenge of larger and richer datasets. GigaScience. 2015;4:s13742-015-0047–0048.

57. Rogers P, Stoner J. Modification of the Sandwich Estimator in Generalized Estimating Equations with Correlated Binary Outcomes in Rare Event and Small Sample Settings. American Journal of Applied Mathematics and Statistics. 2018;3:243–251.

58. R Core Team. R: A Language and Environment for Statistical Computing. Vienna, Austria: R Foundation for Statistical Computing; 2022.

59. Carey VJ. gee: Generalized Estimation Equation Solver. 2022.

60. Nyholt DR. A Simple Correction for Multiple Testing for Single-Nucleotide Polymorphisms in Linkage Disequilibrium with Each Other. Am J Hum Genet. 2004;74:765–769.

61. Xiong Z, Yang F, Li M, Ma Y, Zhao W, Wang G, et al. EWAS Open Platform: integrated data, knowledge and toolkit for epigenome-wide association study. Nucleic Acids Research. 2022;50:D1004–D1009.

62. Sommerer Y, Ohlei O, Dobricic V, Oakley DH, Wesse T, Sedghpour Sabet S, et al. A correlation map of genome-wide DNA methylation patterns between paired human brain and buccal samples. Clinical Epigenetics. 2022;14:139.

63. Koopmans F, Dijkstra AA, Li W-P, Klaassen RV, Gouwenberg Y, Yao S, et al. Human brain prefrontal cortex proteomics identifies compromised energy metabolism and neuronal function in Schizophrenia. In preparation.

64. Langley-Evans SC, McMullen S. Developmental Origins of Adult Disease. Medical Principles and Practice. 2010;19:87–98.

65. Barker DJP. Fetal origins of coronary heart disease. BMJ. 1995;311:171–174.

66. Barker DJP. Fetal origins of cardiovascular disease. Annals of Medicine. 1999;31:3–6.

67. Eaves LJ, Pourcain BSt, Smith GD, York TP, Evans DM. Resolving the Effects of Maternal and Offspring Genotype on Dyadic Outcomes in Genome Wide Complex Trait Analysis (“M-GCTA”). Behav Genet. 2014;44:445–455.

68. Zhang G, Bacelis J, Lengyel C, Teramo K, Hallman M, Helgeland Ø, et al. Assessing the Causal Relationship of Maternal Height on Birth Size and Gestational Age at Birth: A Mendelian Randomization Analysis. PLoS Med. 2015;12:e1001865.

69. Leinonen JT, Pirinen M, Tukiainen T. Disentangling the link between maternal influences on birth weight and disease risk in 36,211 genotyped mother–child pairs. Commun Biol. 2024;7:175.

70. Kato H, Kimura H, Kushima I, Takahashi N, Aleksic B, Ozaki N. The genetic architecture of schizophrenia: review of large-scale genetic studies. J Hum Genet. 2023;68:175–182.

71. Sullivan PF, Yao S, Hjerling-Leffler J. Schizophrenia genomics: genetic complexity and functional insights. Nat Rev Neurosci. 2024;25:611–624.

72. Sekar A, Bialas AR, de Rivera H, Davis A, Hammond TR, Kamitaki N, et al. Schizophrenia risk from complex variation of complement component 4. Nature. 2016;530:177–183.

73. Chindemi G, Abdellah M, Amsalem O, Benavides-Piccione R, Delattre V, Doron M, et al. A calcium-based plasticity model for predicting long-term potentiation and depression in the neocortex. Nat Commun. 2022;13:3038.

74. Sun X, Liu T, Zhao J, Xia H, Xie J, Guo Y, et al. DNA-PK deficiency potentiates cGAS-mediated antiviral innate immunity. Nat Commun. 2020;11:6182.

75. Romero C, Leeuw C de, Schipper M, Maciel B de APC, Heuvel MP van den, Brouwer RM, et al. Genetic overlap between schizophrenia and height implicates pituitary and immune response. 2024:2024.04.10.24305626.

76. Ohlei O, Sommerer Y, Dobricic V, Homann J, Deecke L, Schilling M, et al. Genome-wide QTL mapping across three tissues highlights several Alzheimer’s and Parkinson’s disease loci potentially acting via DNA methylation. 2023:2023.12.22.23300365.

