## Supplementary Notes and Figures for "Intergenerational transmission of complex traits and the offspring methylome"

Supplementary Note 1. Genotyping quality control (QC) and imputation in the Netherlands Twin Register

Genotyping was done according to the manufacturer’s protocols in 37,833 NTR DNA samples from 30,323 individuals, including the families in the current study on the following genotyping platforms: Affymetrix 5-Perlegen (*N* = 1,904), Affymetrix 6.0 (*N* = 10,377), Affymetrix Axiom NL (*N* = 3,536), Illumina Omni 1M (*N* = 445), Illumina Human Quad Array 660W (*N* = 1,501,) and Illumina GSA NTR array (*N* = 20,060). Nearly all the samples, except 44, on the older Illumina Omni 1M, Illumina Human Quad Array 660W, and Affymetrix 5-Perlegen arrays have been re-genotyped on the Illumina GSA NTR array. Therefore – in the end – we only used the Affymetrix Axiom NL, Affymetrix 6.0, and Illumina GSA NTR arrays for imputation. Sample quality control (QC), SNP QC, and data alignment were, however, done on the older arrays as well, because they identify discordant DNA samples, IBD issues, as well as confirm monozygotic (MZ) twins across platforms. Fifteen samples were removed because the DNA of an individual was discordant across the six genotyping platforms and 144 samples were removed because their IBD state did not match the known family structure. For 1,024 MZ twin individuals, we copied the genotype data from twin one to twin two on the Affymetrix 6.0, Affymetrix Axiom NL, and Illumina GSA NTR arrays if they had a different platform genotyped. Thus, all MZ twin pairs have the same genotyping platform. The final Affymetrix Axiom NL array included 534,405 SNPs genotyped for 3,644 individuals, the final Affymetrix 6.0 comprised 537,992 SNPs for 9,049 individuals, and the final Illumina GSA NTR arrays retained 481,898 SNPs for 16,276 individuals.

*Sample QC*

QC of the DNA samples was performed separately by genotype platform. In total 1,439 DNA samples were removed, because their genotyping call rate was below 90%, they had less than 80% available genotypes on each autosomal chromosome and the X chromosome, their reported sex mismatched their genotyped sex, their PLINK heterozygosity F values were outside of -0.10 and 0.10, or their PLINK estimated Identity-By-Descent (IBD) did not match their known family structure. For 1,746 samples, which were genotyped multiple times, one sample was included in further QC steps.

*SNP QC*

SNPs with call rates below 95%, Hardy Weinberg Equilibrium (HWE) p-values below 0.0001, minor allele frequencies (MAF) below 0.01, and Mendelian error rates above 1% were removed from each platform. SNPs were also removed if genotypes differed by more than 1% across plate control samples on the Affymetrix 6.0 (*N* = 4, typed 38-84 times) or Affymetrix Axiom (*N* = 2, typed 33-37 times) platforms or among those samples genotyped multiple times.

*Reference panel alignment and imputation*

We extracted the overlapping SNPs from the 1000 genomes phase 3 (v.5) European populations [1], HRC 1.1 (EGA version) [2, 3], and GONL (v.4) [4] references panels, where the allele frequencies were less than 0.20 apart across all three reference panels. Since the allele frequencies of SNPS can also vary substantially between the reference panels, we further removed SNPs in the NTR, if the NTR allele frequencies were outside of Fr -0.10 and 0.10, where Fr is defined as (*Fr* = (Fmin + Fmax)/2), and Fmin and Fmax are the minimum and maximum frequency, respectively, in any of the 3 references. All palindromic SNPs with allele frequencies of 0.40 to 0.60 were removed. The remaining SNPs were aligned to the positive strand on genome build 37 for all genotyping platforms. The SNP name and reference allele for each of the three genotyping platforms were matched with the HRC 1.1 (EGA version) and GONL reference panels, and then converted to VCF format with PLINK prior to imputation. As the EGA version of the HRC 1.1 does not include GONL, we merged these reference panels following the protocols described elsewhere [2]. Imputation to the combined HRC 1.1 (EGA version) and GONL reference panels was performed in Beagle 5.4 [5] for each of the three remaining genotype platforms in NTR. The platforms were subsequently combined into a single dataset with BCFtools [6], and converted to best guess genotypes with PLINK1.96 [7]. The same approach was used to also impute and combine the data to the 1000 genomes reference data.

**Supplementary Note 2.** Computation of genetic principal components in the Netherlands Twin Register.

Since three genotyping platforms were used in the Netherlands Twin Register, principal components were calculated based on a selection of the remaining genotyped SNPs present in any of the three platforms. However, as not all SNPs were genotyped on all three platforms, these were extracted from the 1000 genomes imputed data.

To create input data for genetic principal component analysis additional SNP filters were employed. SNPs were removed with MAF below 0.05, HWE p-values below 0.001, genotyping call rates below 98%, Mendelian error rates above 1%, imputation info below 90% and in long range linkage disequilibrium (LD) blocks [8]. Pruning was then done to select SNPs in linkage equilibrium (option --indep 50 5 2) in PLINK1.9. The samples with the remaining 110,558 SNPs were extracted from the 1000 genomes reference panel and merged with the same SNP NTR data. We calculated twenty 1000 genomes principal components and projected these on the NTR data with EIGENSTRAT in the smartpca (v.7) software [9].

Supplementary Note 3. Equivalence of the transmitted/non-transmitted alleles design and the genetic parent-offspring model for estimating genotype-environment covariance.

Given phenotypic data in the children and relevant genetic data in the offspring and parents, covariance between additive genetic factors (A) and shared environmental factors (C) [10] can be detected using the ingenious transmitted–non-transmitted alleles design [11–13]. The path diagram associated with this design is depicted in Supplementary Figure 9. In this model, the regression of the offspring phenotype on the genetic values base on the non-transmitted alleles and the transmitted alleles provides the test of genotype-shared environment covariance (cov(AC)). The present aim is to show that the cov(AC) can also be tested by regressing the offspring phenotype on the parental genetic values and offspring genetic values (based on the transmitted alleles). Similarly to Okbay et al. (2022) [14], we show that these two methods are equivalent regarding the test of cov(AC). The two regression models are:

Ph_C_ = b_0_ + b_1_*G{NT}_C_ + b_2_*G{T}_C_ + e	(1)

Ph_C_ = f_0_ + f_1_*(G_M_ + G_F_) + f_2_*G{T}_C_ + e	(2)

where Ph_C_ is the phenotypic score of the child, G{T}_C_ is the genotypic value based on alleles transmitted from the parents to the child, G{NT}_C_ is the genotypic value based on non-transmitted alleles, G_M_ and G_F_ are genotypic values of the mother and father, respectively, e is the residual. We call the model, based on Equation 1, the transmitted/non-transmitted (N/NT) model, and the model, based on Equation 2, the genetic parent-offspring (GPO) model. We show below that the tests of b_1_ = 0 and f_1_ = 0 are equivalent tests of cov(AC).

Given a single diallelic genetic variant (GV) with additive gene action and allele frequency p and q (q=1-p), Supplementary Figure 9 shows the relationship between the alleles and the genotypes in parents and offspring. The variance of the alleles is pq (denoted v in Supplementary Table 9) and the variance of the genotype is 2pq. The parameter *a* represents the direct genetic effect, and the parameter *g* is the source of cov(AC). In Supplementary Figure 9, g is the direct effect of parental phenotype on offspring phenotype. But we assume that the parental phenotypic effect is mediated by the common environment [13, 15], i.e., a latent variable, which is not explicitly modelled here. We consider the covariance matrices of the variables that feature in the T/NT model and GPO model. These are shown in Supplementary Table 9.

Discarding the intercepts, the expression for the regression coefficients is $\Sigma_{X}^{-1}\Sigma_{XY}$, where $\Sigma_{X}$ (2x2) is the covariance matrix of the predictors, and $\Sigma_{XY}$ (2x1) is the matrix of the covariances between the predictors and the dependent variable. Given v=pq, In the N/NT model, we have:

$\Sigma_{XY}$ = 2agv	$\Sigma_{X}^{-1}$ = 1/(2v)	0

2(ag+a)v	0	1/(2v)

and

$\Sigma_{X}^{-1}\Sigma_{XY}$ =	a*g	(b_1_ in Eq 1)

a*g+a	(b_2_ in Eq 2)

In the GPO model, we have

$\Sigma_{XY}$ =	2(agv+(ag+a)v)	$\Sigma_{X}^{-1}$ = 1/(2v)	-1/(2v)

2(ag+a)v	-1/(2v)	1/v

and

$\Sigma_{X}^{-1}\Sigma_{XY}$=	a*g	(f_1_ in Eq 2)

a	(f_2_ in Eq 2).

So, the parameter f_1_ = b_1_ = a*g. The standard errors (s.e.) of the estimates of f_1_ and b_1_ equal s.e.(f_1_) = s.e.(b_1_) = √(1/(2v(N-1))), where N is the sample size (i.e., number of families). Given the equivalence of the T/NT model and the GPO model, either one can be used to test cov(AC) = 0. If genotyping is limited to one parent, it is difficult to determine the transmitted / non-transmitted status of the alleles in the children. In that case, we can still use the GPO model, with a single parent (say, the mother):

Ph_C_ = t_0_ + t_1_*PGS_M_ + t_2_*PGS{T}_C_ + ε	(3)

The test of cov(AC) is t_1_=0. The parameters t_1_ and t_2_ are:

$\Sigma_{XY}$ =	agv(ag+a)v	$\Sigma_{X}^{-1}$ =	2/(3v)	-1/(3v)

2(ag+a)v	-1/(3v)	2/(3v)

and

$\Sigma_{X}^{-1}\Sigma_{XY}$ =	(2*a*g)/3	(t_1_ in Eq 3)

(2*a*g)/3+a	(t_2_ in Eq 3).

The standard error of t_1_, s.e.(t_1_), equals √(2/(3v(N-1)). As expected, given the difference in the information, the power to reject f_1_=0 or b_1_=0 is greater than the power to reject t_1_=0: the test statistic f_1_^2^/s.e.(f_1_)^2^ is asymptotically 3 times larger than the test statistic t^12^/s.e.(t_1_)^2^.

In considering these tests, we assumed that the direct genetic effect is identical in the parents and the offspring (parameter a in Supplementary Figure 9). This assumption can be relaxed. Given direct genetic effects a_P_ and a_C_ in parents and offspring, respectively, we have:

b_1_=a_P_*g and b_2_=a_P_*g + a_C_	(T/NT model)

f_1_= a_P_*g and f_2_=a_C_	(GPO model, 2 parents)

t_1_=(2*a_P_*g)/3 and t_2_=(2*a_P_*g)/3+a_C_	(GPO model, 1 parent)

*Simulations*

We simulated phenotypes in the offspring and transmitted and non-transmitted polygenic scores for 100,000 parent-offspring trios based on the relation in the path diagram in Supplementary Figure 9. These simulations empirically demonstrate the equivalence between the T/NT model (equation 1) and the GPO model (equation 2) for detecting cov(AC) (Supplementary Table 10).

*Conclusion*

In conclusion, the GPO model and the T/NT model provide equivalent tests of cov(AC). Arguably, the GPO model is preferable for two reasons. One, it does not require one to determine the T/NT status of the parental alleles, which can be effortful. Two, it allows one to test cov(AC), even if the parental genetic data is limited to one parent. We have considered a single locus, but the equivalence generalizes to polygenic scores (PGS), based on multiple associated loci. Specifically, substituting a polygenic score for the single GV in Supplementary Figure 9 would not alter the present result concerning the equivalence of the parameters f_1_ (Eq. 2) and b_1_ (Eq. 1). The only difference is that the parameter v (presently v=pq) would equal ½σ^2^_PGS_, i.e., half of the additive genetic variance attributable to the PGS.

Supplementary Figure 1. Variability of DNA methylation probes per decile.

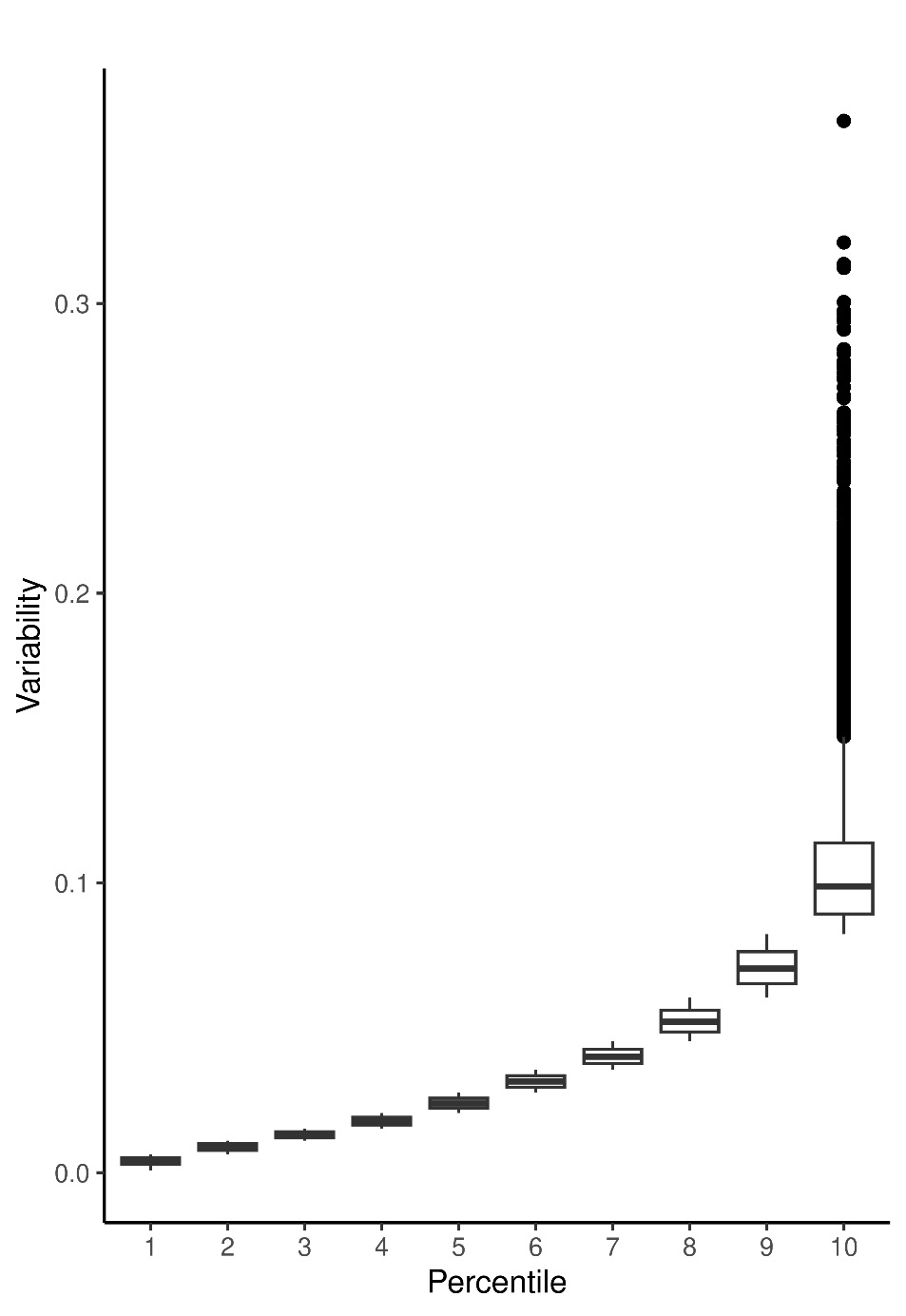

Supplementary Figure 2. Manhattan plots of the association results for the offspring and parental PGSs for schizophrenia with the 72,697 DNA methylation probes as assessed when simultaneously modelling all PGSs. The red lines indicate the Bonferroni significance threshold corrected for the number of independent CpGs (*p* = 2.7 × 10^−5^).

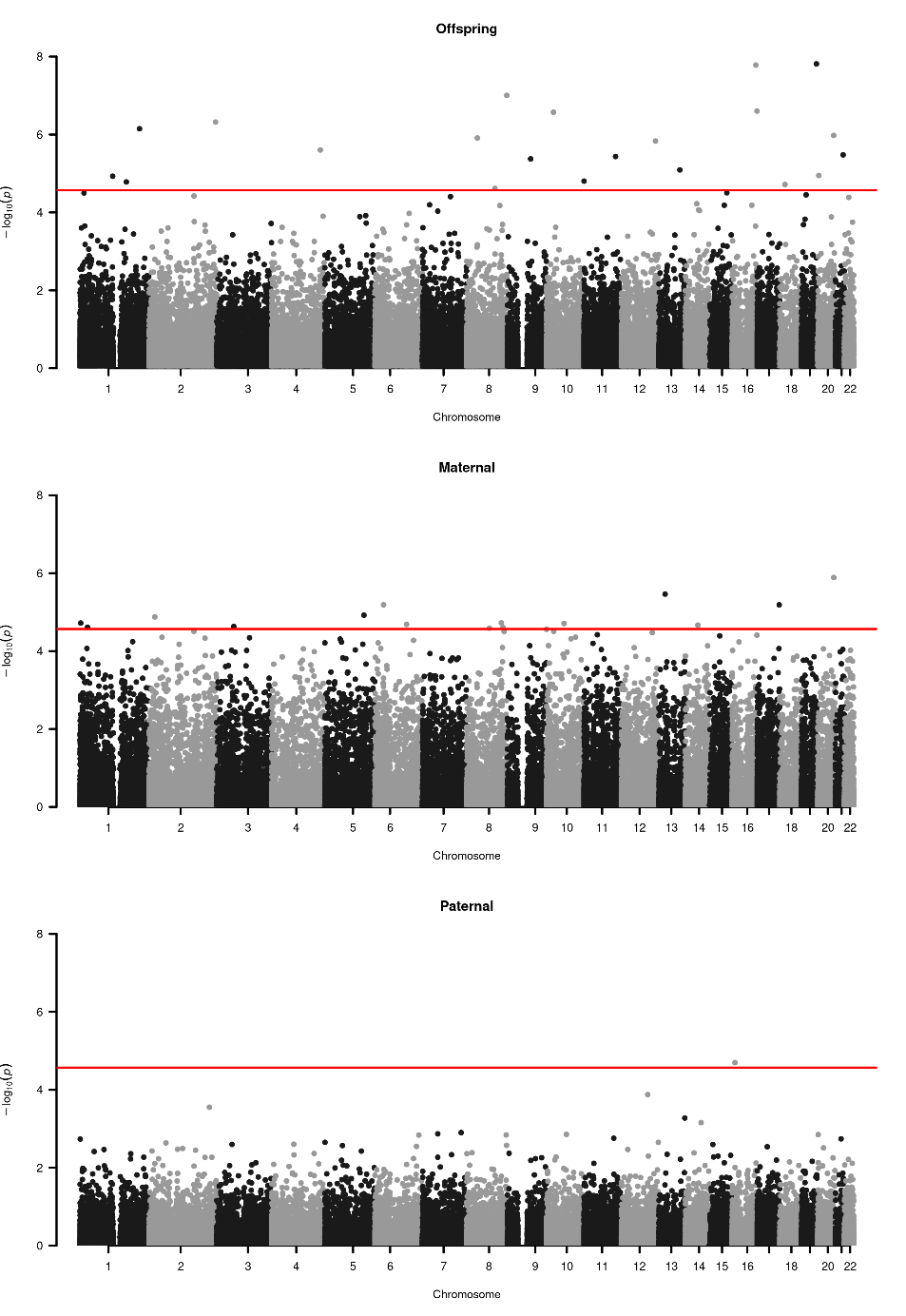

Supplementary Figure 3. Manhattan plots of the association results for the offspring and parental PGSs for smoking initiation with the 72,697 DNA methylation probes as assessed when simultaneously modelling all PGSs. None of the CpGs reached the Bonferroni significance threshold corrected for the number of independent CpGs (*p* = 2.7 × 10^−5^)

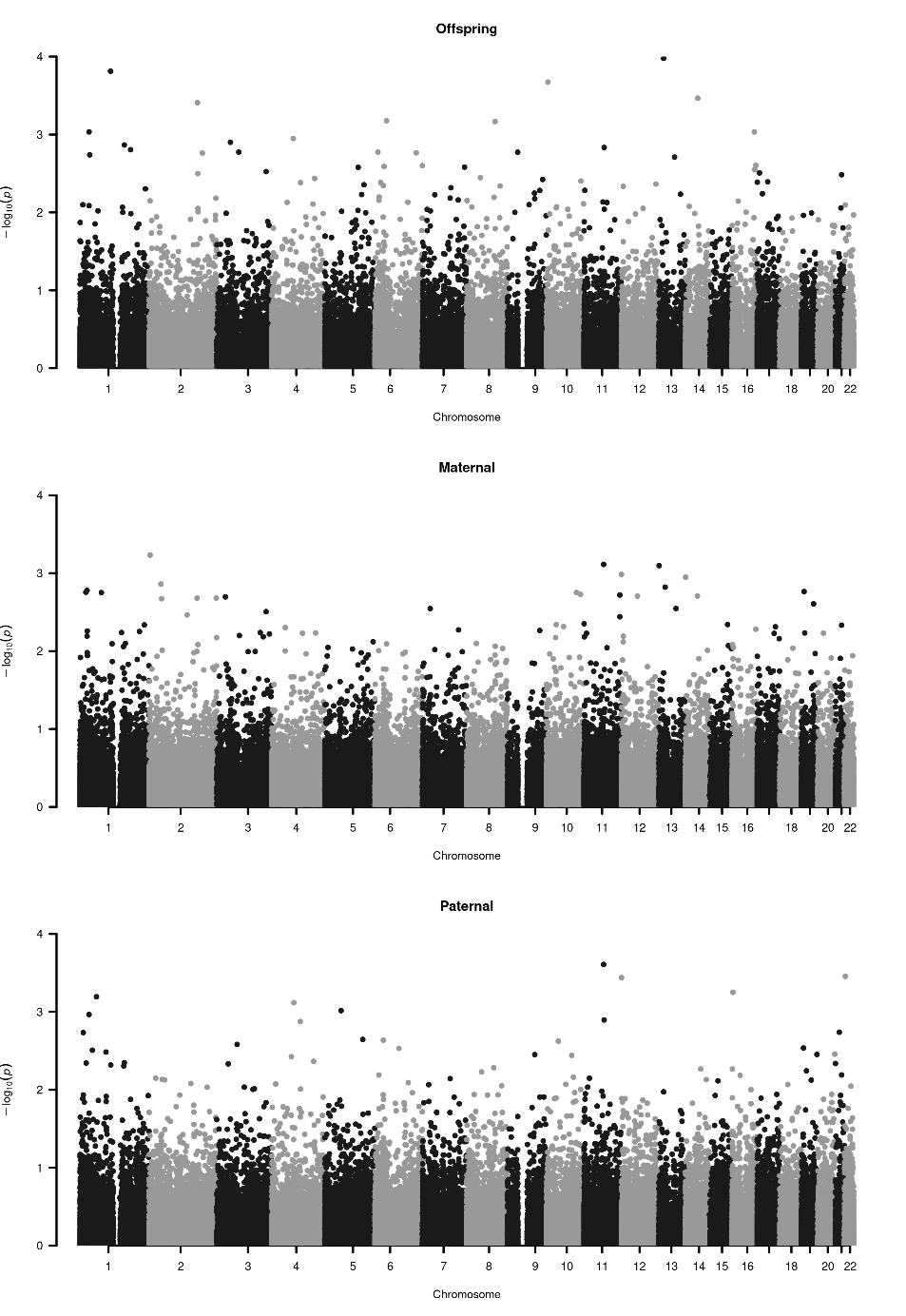

Supplementary Figure 4. Manhattan plots of the association results for the offspring and parental PGSs for educational attainment with the 72,697 DNA methylation probes as assessed when simultaneously modelling all PGSs. The red lines indicate the Bonferroni significance threshold corrected for the number of independent CpGs (*p* = 2.7 × 10^−5^).

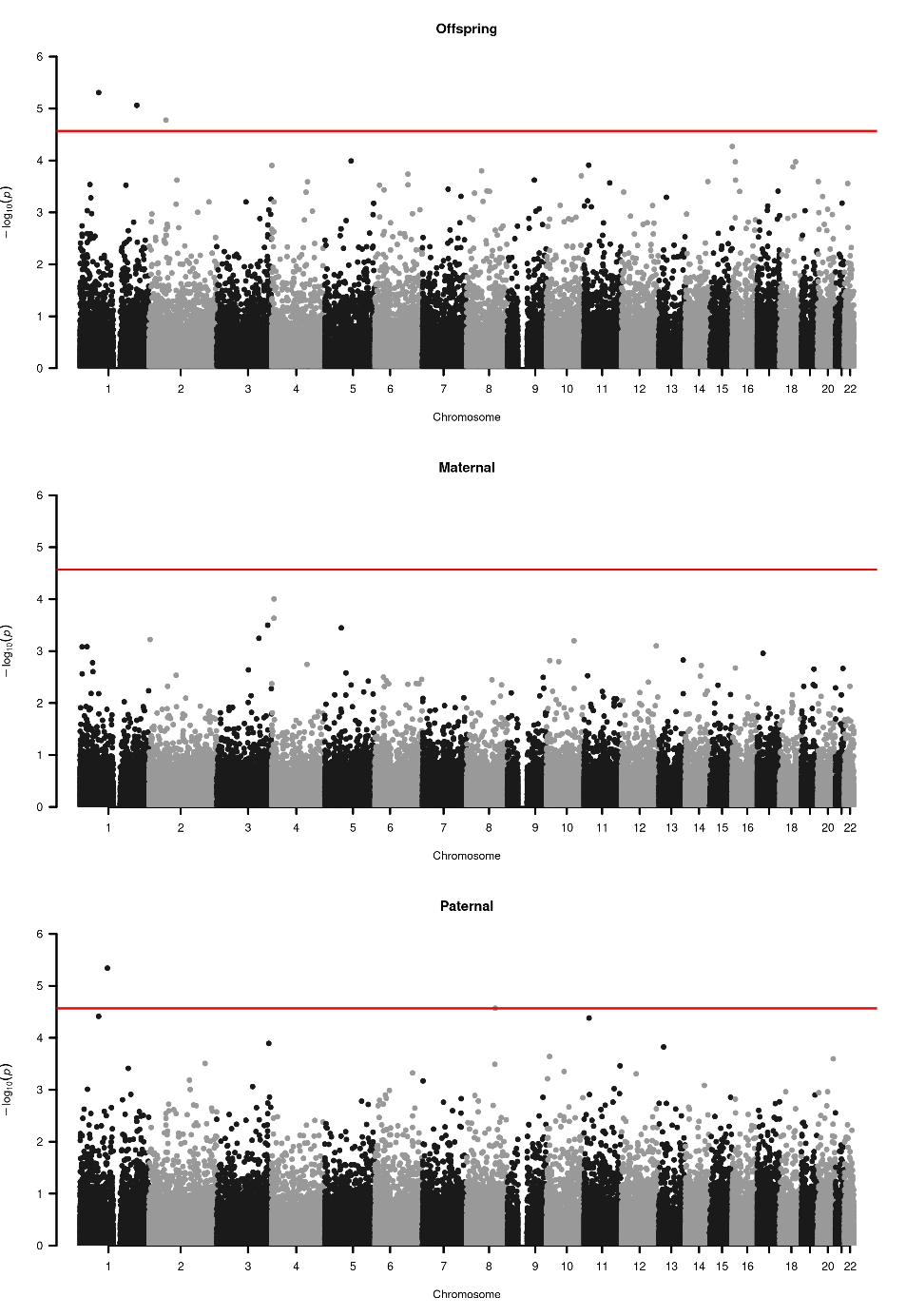

Supplementary Figure 5. Manhattan plots of the association results for the offspring and parental PGSs for social deprivation with the 72,697 DNA methylation probes as assessed when simultaneously modelling all PGSs. The red lines indicate the Bonferroni significance threshold corrected for the number of independent CpGs (*p* = 2.7 × 10^−5^).

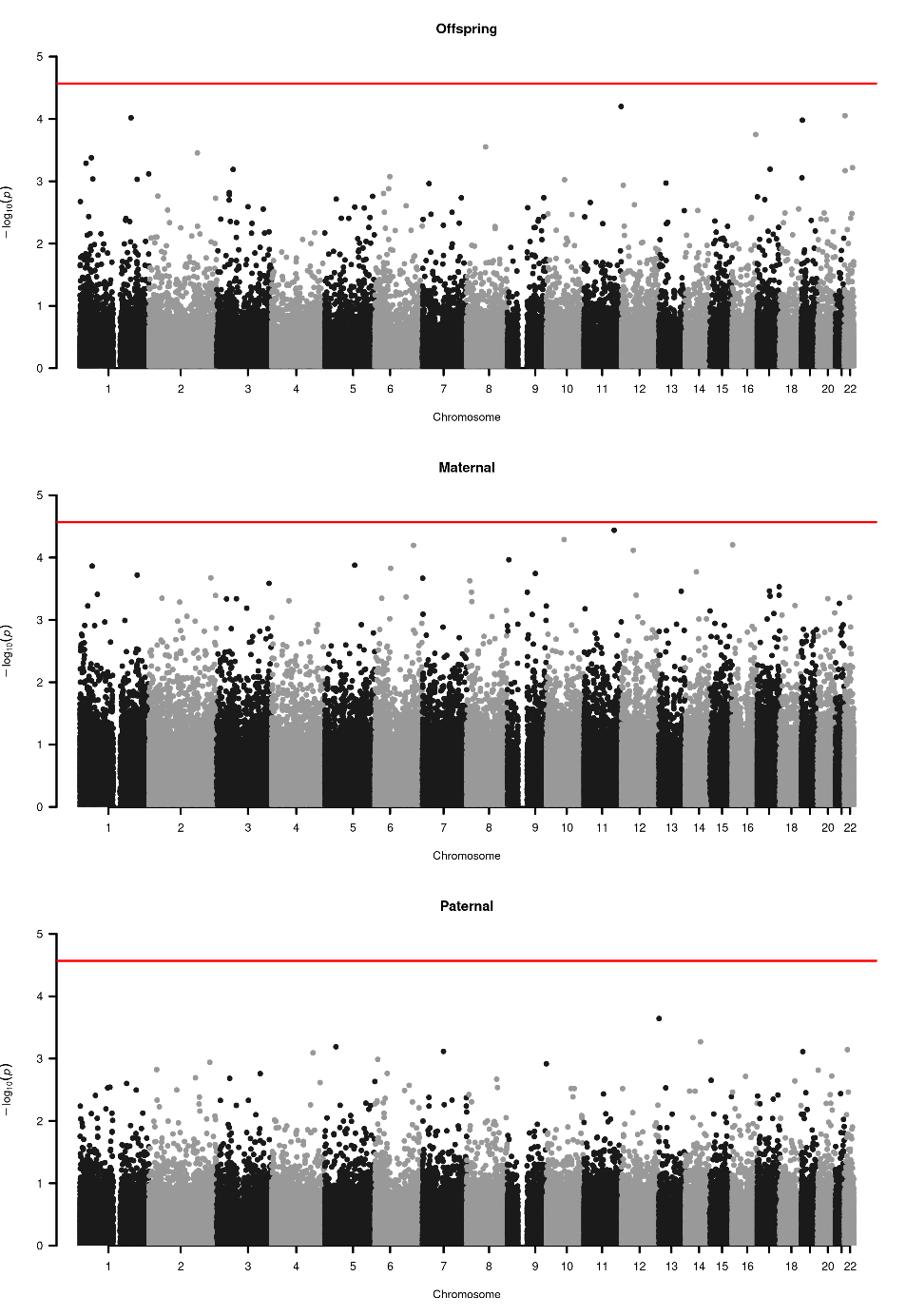

Supplementary Figure 6. Manhattan plots of the association results for the offspring and parental PGSs for BMI with the 72,697 DNA methylation probes as assessed when simultaneously modelling all PGSs. The red lines indicate the Bonferroni significance threshold corrected for the number of independent CpGs (*p* = 2.7 × 10^−5^).

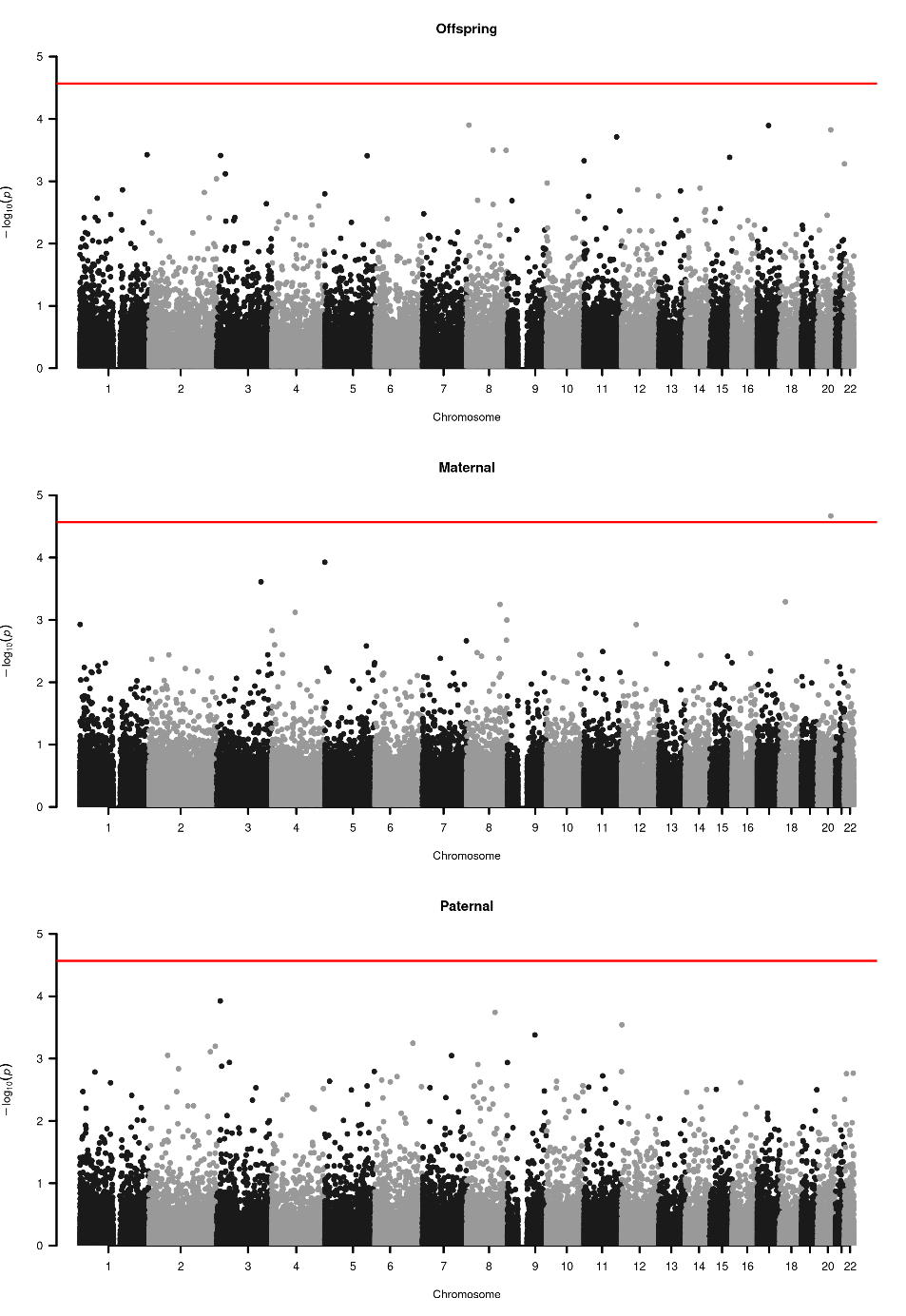

Supplementary Figure 7. Manhattan plots of the association results for the offspring and parental PGSs for height with the 72,697 DNA methylation probes as assessed when simultaneously modelling all PGSs. The red lines indicate the Bonferroni significance threshold corrected for the number of independent CpGs (*p* = 2.7 × 10^−5^).

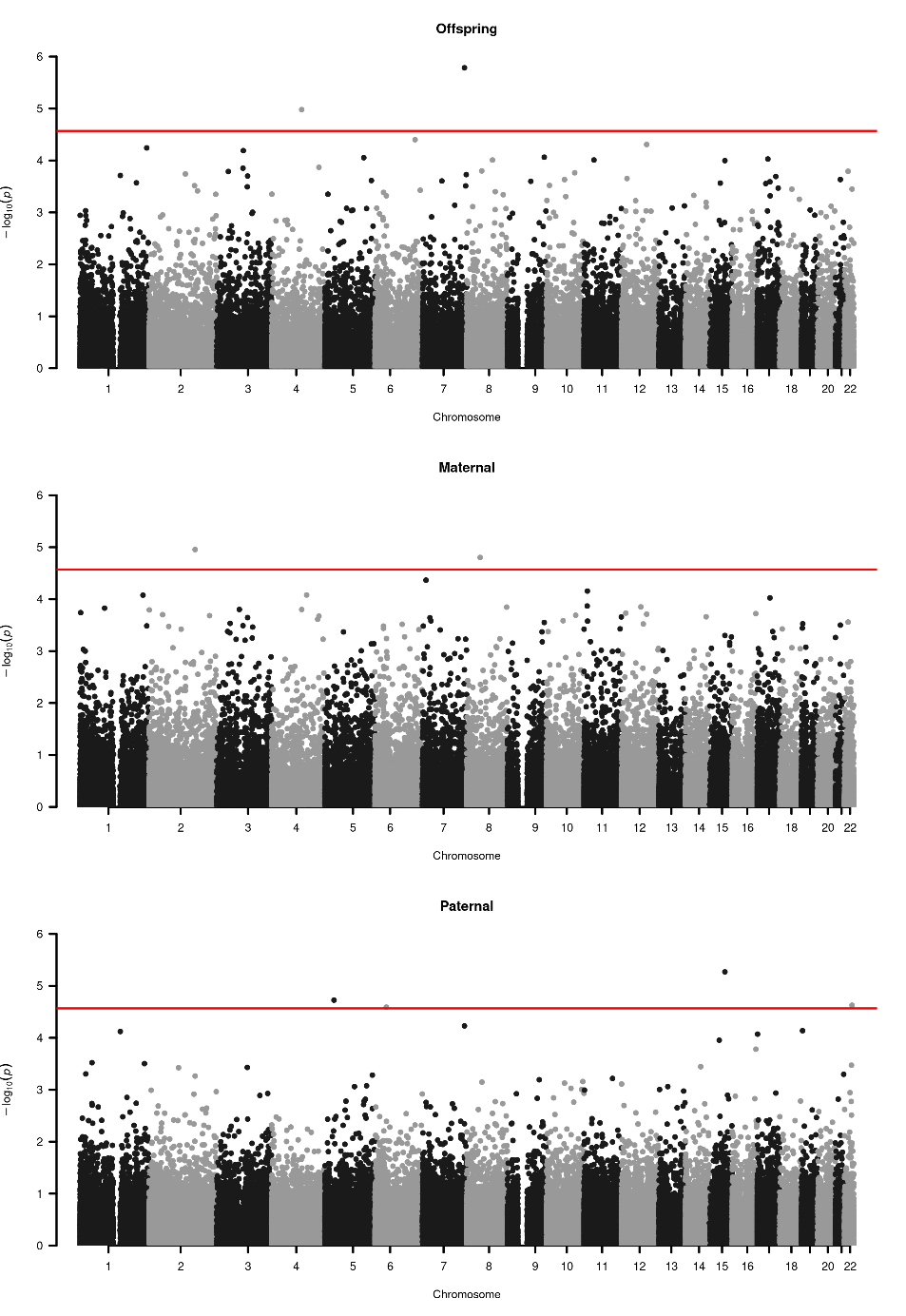

**Supplementary Figure 8.** Venn diagram of the overlap between blood *cis* methylation quantitative trait loci (mQTLs) [16] associated with the significantly associated (*α* = 2.7 × 10^−5^) CpG cg01633359 for the paternal polygenic score for height **(a)** and with the significantly associated CpG cg03480605 for the offspring polygenic score for height **(b)** and genome-wide significant (*p* < 5 x 10^-8^) SNPs for height [17].

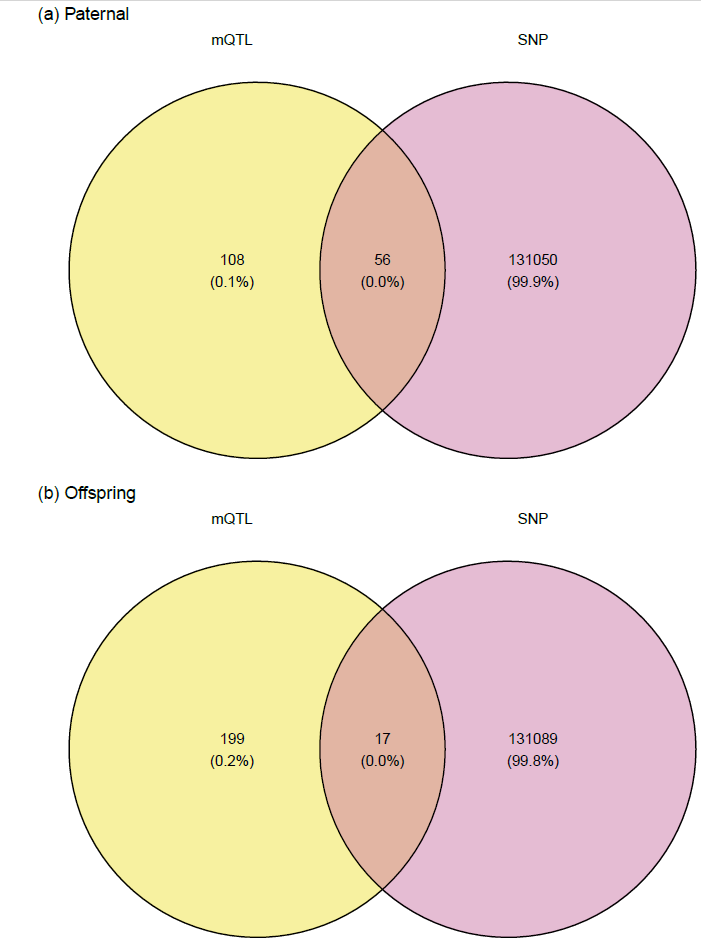

**Supplementary Figure 9.** Transmitted/non-transmitted design. The parameter *a* represent the direct genetic effect on the phenotype. The path from parental phenotype to the child’s phenotype, with path coefficient *g*, is the source of genotype-shared environment covariance (cov(AC)). In theory, this relationship is mediated by the children’s shared environment [10, 15].

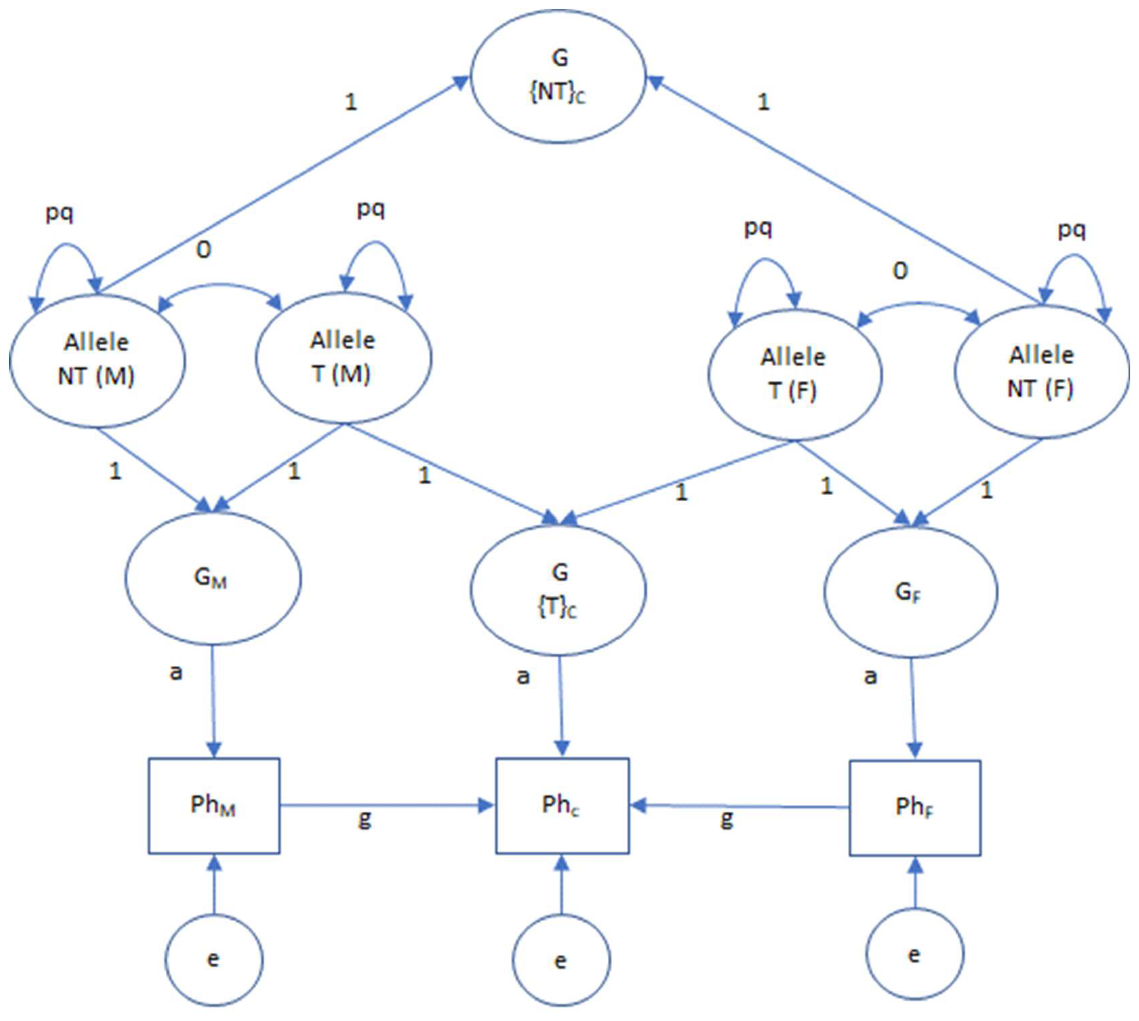

Supplementary references

1.	Auton A, Abecasis GR, Altshuler DM, Durbin RM, Abecasis GR, Bentley DR, et al. A global reference for human genetic variation. Nature. 2015;526:68–74.

2.	McCarthy S, Das S, Kretzschmar W, Delaneau O, Wood AR, Teumer A, et al. A reference panel of 64,976 haplotypes for genotype imputation. Nat Genet. 2016;48:1279–1283.

3.	The Haplotype Reference Consortium - EGA European Genome-Phenome Archive. https://ega-archive.org/studies/EGAS00001001710. Accessed 6 April 2023.

4.	Boomsma DI, Wijmenga C, Slagboom EP, Swertz MA, Karssen LC, Abdellaoui A, et al. The Genome of the Netherlands: design, and project goals. Eur J Hum Genet. 2014;22:221–227.

5.	Browning BL, Tian X, Zhou Y, Browning SR. Fast two-stage phasing of large-scale sequence data. The American Journal of Human Genetics. 2021;108:1880–1890.

6.	Li H. A statistical framework for SNP calling, mutation discovery, association mapping and population genetical parameter estimation from sequencing data. Bioinformatics. 2011;27:2987–2993.

7.	Chang CC, Chow CC, Tellier LC, Vattikuti S, Purcell SM, Lee JJ. Second-generation PLINK: rising to the challenge of larger and richer datasets. GigaScience. 2015;4:s13742-015-0047–0048.

8.	Abdellaoui A, Hottenga J-J, Knijff P de, Nivard MG, Xiao X, Scheet P, et al. Population structure, migration, and diversifying selection in the Netherlands. Eur J Hum Genet. 2013;21:1277–1285.

9.	Price AL, Patterson NJ, Plenge RM, Weinblatt ME, Shadick NA, Reich D. Principal components analysis corrects for stratification in genome-wide association studies. Nat Genet. 2006;38:904–909.

10.	Fulker, D. W. Genetic and cultural transmission in human behavior. Proceedings of the Second International Conference on Quantitative Genetics, Sinauer Associates Inc.; 1988. p. 318–340.

11.	Bates TC, Maher BS, Medland SE, McAloney K, Wright MJ, Hansell NK, et al. The Nature of Nurture: Using a Virtual-Parent Design to Test Parenting Effects on Children’s Educational Attainment in Genotyped Families. Twin Research and Human Genetics. 2018;21:73–83.

12.	Kong A, Thorleifsson G, Frigge ML, Vilhjalmsson BJ, Young AI, Thorgeirsson TE, et al. The nature of nurture: Effects of parental genotypes. Science. 2018;359:424–428.

13.	Balbona JV, Kim Y, Keller MC. Estimation of Parental Effects Using Polygenic Scores. Behav Genet. 2021;51:264–278.

14.	Okbay A, Wu Y, Wang N, Jayashankar H, Bennett M, Nehzati SM, et al. Polygenic prediction of educational attainment within and between families from genome-wide association analyses in 3 million individuals. Nat Genet. 2022;54:437–449.

15.	Keller MC, Medland SE, Duncan LE, Hatemi PK, Neale MC, Maes HHM, et al. Modeling Extended Twin Family Data I: Description of the Cascade Model. Twin Research and Human Genetics. 2009;12:8–18.

16.	Min JL, Hemani G, Hannon E, Dekkers KF, Castillo-Fernandez J, Luijk R, et al. Genomic and phenotypic insights from an atlas of genetic effects on DNA methylation. Nat Genet. 2021;53:1311–1321.

17.	Yengo L, Sidorenko J, Kemper KE, Zheng Z, Wood AR, Weedon MN, et al. Meta-analysis of genome-wide association studies for height and body mass index in ∼700000 individuals of European ancestry. Hum Mol Genet. 2018;27:3641–3649.
